# Genomic and Functional studies identify *RPSA* as a risk gene for ALS and other neurological diseases

**DOI:** 10.1101/2025.07.28.25332301

**Authors:** Xiaoyu Qian, Brett W Stringer, Cheong W. Wong, Ang Li, Victoria Sjalim, Fei-Fei Cheng, Mike J. Thompson, Ruolan Zhao, Tian Lin, Anjali K. Henders, Pamela A. McCombe, Naomi R. Wray, Allan F. McRae, Jean Giacomotto, Fleur C. Garton

## Abstract

Amyotrophic lateral sclerosis (ALS) is a progressive neurodegenerative disease characterised by motor neuron deterioration. Genetic factors play a significant role in all cases, with 15 genome-wide significant study (GWAS) risk loci identified to date. Follow-up of these loci is a powerful strategy for research translation, as drug targets supported by genetic evidence are more likely to succeed in clinical development.

Here, we focus on the *RPSA-MOBP* locus on chromosome 3 (lead SNP, rs631312, OR = 1.08 95% CI: 1.06–1.10, p = 3.3 × 10⁻¹²). We employ integrative *in silico* analyses to prioritise candidate genes, combining multiple ‘omics-based approaches, including Functional Mapping and Annotation (FUMA), Polygenic Priority Scoring (PoPS), Transcriptome-Wide Association across/within tissues (TWAS), gene-based test (mBAT-combo), chromatin interaction mapping (H-MAGMA), and Summary data Mendelian Randomisation (SMR), with GWAS data (*N*_cases_ = 29,612, *N*_controls_ = 122,656).

Both *RPSA* and *MOBP* were prioritised as candidate genes in multiple analyses. *In-vivo* expression analyses in ALS blood or iPSC-motor neurons were unremarkable for these genes but also other-relevant ALS genes. *RPSA*, highly conserved in zebrafish (92% homology), was selected for functional modelling, noting previously generated *Mobp*-ko mice show minimal phenotypic changes. CRISPR/Cas9-induced *rpsa* loss-of-function (LOF) in zebrafish triggers progressive and severe phenotypes mimicking pathology observed in SMN- and TDP43-deficient zebrafish, two key proteins/genes associated with diseases of the motor neurons. *RPSA*-deficient animals exhibit marked motor neuron axon pathology, progressive loss of motor function and rapid decline culminating with premature death at around 7 days-post-fertilisation. These phenotypes were notably similar to those observed in SMN and TDP-43 zebrafish models, together with prominent cardiovascular abnormalities.

This study identifies *RPSA* as a critical gene for motor neuron health, with implications for ALS pathogenesis. The *RPSA/MOBP* locus is also associated with other neurodegenerative diseases including frontotemporal dementia/FTD, corticobasal degeneration/CBD and progressive supranuclear palsy/PSP, highlighting its potential as a therapeutic target for multiple conditions.

## Introduction

Amyotrophic lateral sclerosis (ALS) is a neurodegenerative disorder with a lifetime risk of ∼1 in 350^1^. It is the most common form of motor neuron disease, usually arising in adulthood between the ages of 50-75, noting earlier onset can occur. Presentation is heterogeneous, affecting both motor and extramotor domains, with cognitive, emotional, and behavioural changes that can overlap with frontotemporal dementia^1^. Approved drugs (variable worldwide) currently include Riluzole^2^ (glutamate antagonist), Endaravone^3^ (antioxidant) and Qalsody^4^ (an antisense oligonucleotide targeting *SOD1* mutation (∼1-2% of all ALS cases)). Both Riluzole and Edaravone can slow disease progression to some extent but do not change the typical (fatal) trajectory. With limited therapeutic options, ventilatory support is currently the most significant intervention that can prolong life, in a disease which is otherwise fatal within ∼3-5 years^1^.

About 10% (Asian-ancestry), to 15% (European-ancestry) of ALS cases have a Mendelian form of disease and carry a pathogenic variant in a known ALS gene^5^. Investigation of disease mechanisms and testing of treatments have focussed on these Mendelian forms of ALS, using e.g. *SOD1* or *C9orf72* mutant models to recapitulate mechanisms/intervention responses. However, ALS disease aetiology is considered complex in most cases, involving both genetic and environmental factors. Now emerging are investigations on the more common (and complex) presentation, ∼85-90% cases, who are expected to carry a portfolio of risk variants^6–10^. Notably, the heritability of ALS, and the estimated genetic contribution, is large (∼43%, 95% CI, 0.34–0.53)^11^. In contrast to biomarkers which could reflect both state and trait, genetic risk factors from GWAS can point to causal mechanisms. Follow-up of GWAS risk loci^7,8,12^ are now increasingly being utilised in drug development. This evidence is considered a valuable research translation strategy given genetically supported drug targets (such as from GWAS) are twice as likely to make it through the drug pipeline than targets without genetic support^13^. Given the extremely high failure rate of clinical trials (including for ALS), placing resources into the follow-up of GWAS findings is incredibly important for the affected community.

In ALS, 15 GWAS risk loci have been identified to-date (most recent cohort N_cases_=29,612, N_controls_=122,656)^2^, with many more expected as sample sizes increase. One of these loci is in the *RPSA/MOBP* region on chromosome 3. The *RPSA-MOBP* locus association with ALS was first identified in 2016^10^ and remains significant in the most recent GWAS, with a consistent signal across ancestries (European, Beta effect= 0.079 (0.012)) and non-European/Asian ancestry, Beta effect= 0.084 (0.036)) analyses, (meta p value= 3.3 × 10^−12^))^2^.

Interestingly, this same locus (*RPSA-MOBP* on chromosome 3) has been associated with traits from brain imaging data, such as cortical thickness^14^ and other, clinically overlapping (but generally rarer) neurodegenerative conditions. This includes frontotemporal dementia/FTD (N_cases_=4,685, N_controls_=15,308)^15^, corticobasal degeneration/CBD (N_cases_=219, N_controls_=3,750)^16^ and progressive supranuclear palsy/PSP (N_cases_=2,779, N_controls_=5,584)^17^. For ALS, the top hit is rs631312, (OR=1.083, 95% CI: 1.06–1.10, P value=3.3 × 10^−12^)^2^, for PSP it is the same top SNP rs631312 (r^2^=1, OR=1.46, 95% CI: 1.38–1.54, 4.60 × 10^−20^)^17^ for FTD rs1009966 (r^2^=0.31, OR= 1.16, 95% CI: 1.11–1.21, P value=2.36 × 10^−8^)^15^, and for CBD it is rs1768208, r^2^=0.99, OR=1.71, 95% CI: 1.36–2.15, P value=2.07 × 10^−7^)^16^. In PSP, *MOBP* has been nominated as the causal gene in the locus^17^. This is based on evidence from oligodendrocyte single cell data and downregulation in PSP brains, in line with its role in synthesis and maintenance of myelin. While in ALS, both *MOBP* and *RPSA* have been linked using SMR approach with human expression data^2^.

Here, we carry out a comprehensive functional analysis of this locus to determine potential mechanisms of this association. We focus on using the most well powered GWAS cohort, ALS and controls, using *in-silico* (mapping approaches to fine-map causal loci and genes using summary statistics and expression data), and *in-vivo* (zebrafish-CRISPR) approaches. This includes colocalisation analyses with other conditions to determine if results (and subsequent therapeutic target) could be relevant/applied in the context of multiple neurological diseases.

## Methods

### Integrative annotation of the ALS genome-wide association data

The most recent, and largest publicly available ALS GWAS summary data from van Rheenen et al.^2^ was used to perform all *in-silico* analyses (N_cases_=29,612, N_controls_=122,656). Briefly, 15 independent loci reached genome-wide significance in the European-ancestry analysis (P < 5.0 × 10⁻⁸). Meta-analyses with a smaller, independent Asian-ancestry cohort replicated 12 loci. In these regions, either the top European SNP or a high-LD proxy (r² = 0.99) was present. For the remaining three loci, the lead European SNPs were not found in the Asian-ancestry cohort, likely due to their low minor allele frequency (MAF 0.6–1.6%)). The *RPSA/MOBP* locus on chromosome 3 locus was one of the 12 replicated loci with a consistent effect size estimated in both European and cross-ancestry results. As such we have run either European only, and/or cross-ancestry (European and Asian-ancestry) summary statistics for follow-up analyses below.

### Fine Mapping the ALS GWAS loci

We applied three fine-mapping tools; SuSiE^18^, MsCAVIAR^19^ and FINEMAP^20^, to estimate variant level posterior probabilities of causality and to define credible sets likely to contain causal variants. Each of these tools was run using standardised summary statistics and locus-specific LD matrices (1000 Genomes Phase 3 European panel or UK Biobank 20K reference), with default or recommended settings unless otherwise specified.

Briefly FINEMAP^20^ is a Bayesian method that evaluates different configurations of causal variants in each locus. It uses a shotgun stochastic search (SSS) to provide Posterior Inclusion Probabilities (PIPs) and 95% credible sets for causal inference. SSS is efficient as it explores the configuration space by randomly sampling configurations, giving higher priority to those with greater posterior probabilities. The later developed SuSiE^18^ (Sum of Single Effects) method that uses a Bayesian regression framework to model multiple causal variants per region. Credible sets with high coverage and low redundancy are generated by estimating the PIP for each SNP. MsCAVIAR^19^ (multi-study CAVIAR) jointly analyses multiple GWAS datasets or ancestries, accounting for LD differences and improving cross-population resolution. It assigns causal probabilities to SNPs under the assumption of a shared causal architecture and employs an exhaustive search that evaluates all possible causal configurations within a given region.

### Functional annotation and mapping

We used FUMA (v1.3.7) to functionally annotate ALS GWAS summary statistics by mapping genes and biological pathways. SNPs are mapped to genes based on position, expression quantitative trait loci (eQTL) mapping, and chromatin interaction (MAGMA = v1.08) using sources including GTEx (v8), Roadmap Epigenomics, and ENCODE. We used the 1KG/Phase3 reference panel and other default parameters for both European and cross-ancestry ALS GWAS results. Briefly, independent significant SNPs were defined with a P-value threshold of <5×10^−8^, and a linkage disequilibrium (LD) r^2^ of <0.6. Within this pool of independent SNPs, lead SNPs were defined as those most highly associated and independent from other lead SNPs (LD of r^2^<0.1). The lead SNPs and those in LD with them were annotated as risk loci (a 250kb window, r^2^ ≥ 0.6).

Significant genes, based on gene set from Ensemble v92, were defined using a Bonferroni-adjusted P-value (0.05/19,290) P < 2.62 × 10^−6^.

### Gene-based association testing

Gene-based association tests (mBAT-combo^21^, MBAT and fastBAT^22^) were used to test the aggregated effect of the GWAS variants within each locus to identify and prioritise ALS-associated genes, noting that methods are similar but mBAT-combo is the most powerful. For this analysis, LD estimates were based on UKB imputed dataset (n=20,000 unrelated individuals) with a 50kb flanking window. Briefly, fastBAT tests sum chi-squared statistics (i.e. squared z-statistics) with appropriate weighting and statistical significance is considered with Bonferroni/FDR. MBAT-combo typically improves power as it combines fastBAT and mBAT, which can handle ‘masking effects’ (i.e., where the product of the effects of two variant alleles is in the opposite direction to their LD correlation).

### Transcriptome-wide association testing

Identification of gene expression-trait associations (Transcription-wide association (TWAS)) was carried out using publicly available eQTL reference data and GWAS summary statistics. We applied a suite of TWAS approaches, given each leverages distinct modelling frameworks and assumptions, despite using similar resources.

Firstly, we applied the widely used TWAS-FUSION^23^. This has a framework that models genetically predicted expression on a tissue-by-tissue basis, utilising various expression reference panels and methods to account for different genetic architectures. Secondly we used UTMOST (Unified Test for MOlecular SignaTures).^24^ This extends FUSION by performing a unified test across multiple tissues, to capture tissue-specific genetic effects and provide a more comprehensive view of the gene expression landscape. SPrediXcan^25^ (Summary-PrediXcan) uses precomputed gene expression prediction models to run in a tissue-by-tissue manner, to estimate the association between genetically predicted gene expression and ALS (but does not explicitly account for correlation between tissues). TF-TWAS^26^ (Transcription Factor TWAS) extends traditional TWAS by incorporating transcription factor binding site information to model gene regulation, thereby enhancing biological interpretability and the ability to detect regulatory effects. GIFT (Gene-level Inference of Functional Traits)^27^ provides a probabilistic gene-trait association estimates by integrating TWAS with fine-mapping and uncertainty in the expression prediction weights and association statistics.

Finally, we used the recently developed TWAS-CONTENT^28^ approach to detect candidate genes using different expression models. This accounts for multiple testing across contexts to improve power and generate context-specific and context-shared components of expression. We used two strategies, one running CONTENT weights alone (generated using an elastic net approach on European individuals’ tissue-shared gene expression data (GTEx)) and another with the classic tissue-by-tissue approach (where the weights are tissue specific). Hierarchical FDR (Benjamini-Hochberg false discovery rate) was used to control the FDR within and across contexts (rather than adjusting it globally across all contexts at once to preserve power in smaller but critical tissue cohorts and avoid bias from context imbalance^28^).

### Polygenic Priority Scoring of the ALS GWAS (PoPS)

The similarity-based gene prioritisation method, Polygenic Priority Score (PoPS)^29^ was used to identify and rank candidate genes. PoPS integrates GWAS summary statistics with diverse functional genomic features—including gene expression, protein-protein interactions, and pathway annotations (many of which are not jointly considered in other tools)—to highlight genes enriched for trait-relevant biological signatures. Using these predefined gene features, PoPS fits a linear model to assign each gene a priority score based on the similarity of its functional profile to the genome-wide polygenic signal. Joint enrichment is estimated via a generalised least squares regression model which also includes a matrix of gene-level covariates such as gene length. The method was run using recommended default parameters, with gene-level outputs ranked by their PoP Score.

### Chromatin-informed mapping of genes in Brain (H-MAGMA)

H-MAGMA (v1.10) was used to map SNPs to genes by integrating ALS GWAS summary statistics with 3D chromatin interaction data from adult brain tissue. Unlike standard MAGMA, which relies solely on linear genomic proximity, H-MAGMA incorporates Hi-C chromatin interaction maps to link non-coding variants to their target genes based on 3D genome architecture. Gene annotations were generated using MAGMA^30^ based on the GRCh37/hg19 reference genome and SNP locations from the ALS GWAS dataset.

Precomputed Hi-C interaction data enabled resolution of distal enhancer-promoter loops, improving the identification of regulatory effects. This process mapped 18,266 SNPs to genes, enhancing detection of functionally relevant loci. SNP-wise gene analysis was conducted using multiple regression models that accounted for linkage disequilibrium, using the 1000 Genomes Project European reference panel. SNP p-values were transformed into Z-scores, and significant genes were identified following false discovery rate (FDR) correction. All genome and gene location files were sourced from the CNCR MAGMA repository.

### Summary-data based Mendelian Randomisation with xQTL data

We applied Summary-data based Mendelian Randomisation (SMR) to integrate ALS GWAS data with xQTL datasets (expression, splicing, methylation and protein QTLs) from brain and blood^31^. The goal of SMR is to test whether the association between a gene and disease is driven by a shared causal variant- providing statistical evidence that a specific SNP may be responsible for both. This complements TWAS but evaluates causality rather than correlation, to support taking SNP findings forward into a costly downstream analysis.

We used large-scale QTL resources, including eQTLgen (Blood, N=31,684), BrainMeta (Brain, N=2,865, expression QTL^32^, splice QTL^32^), Interval^33^, Fenland^34^, SCALLOP^35^ (plasma protein QTLs n=10,708, n=3,301, and N=30,931) with methylation from BrainMeta (Brain, N=1,160^32^) and Blood (LBC+BSGS, N=1,980)^36^. While previous SMR had been run using eQTL (MetaBrain, eQTLGen Consortium) and mQTL (prefrontal cortex, whole blood mQTL datasets)^2^ to prioritise genes (i.e. *CCR8, RPSA, MOBP* (brain expression) *RPSA, MOBP, SNORA6A2* (blood methylation)) no HEIDI test was applied,^2^ making it unclear if whether associations reflected pleiotropy or linkage.

In this analysis, we apply both a genome-wide significance threshold and a regional threshold specific to chromosome 3 (number of genes in the locus (p<0.05/13)), requiring all reported associations to also pass the HEIDI test (p > 0.05). This allowed us to prioritise candidate genes and SNPs with stronger evidence of shared genetic causality with ALS for functional validation.

### *In-vivo* human ALS expression data

To examine changes in transcriptomic expression in disease, blood (N=174, *N_cases_*= 121, *N_controls_* = 53)^37^ and iPSC-MN (N=255, *N_cases_*=213, *N_controls_*=42) RNAseq data^38^ were analysed. The blood samples were from Australian ALS cases and controls (collected with written consent (2012-2022) and study approval (Metro North Health Human Research Ethics Committee EC00172 (2006/047) and the University of Queensland (2021/HE002682)). Clinical data were recorded (research nurses/neurologists) on a secure server that included the generation of a de-identified subject ID. ALS cases fulfilled the revised El Escorial criteria for possible, probable (lab-supported) or definite ALS. Control subjects were unrelated, age-matched individuals free of neuromuscular diseases, recruited as either partners or friends of patients with ALS or community volunteers. Blood samples were collected in PAXgene tubes, RNA extracted (PAXgene Blood miRNA Kit, QIAcube robot) libraries prepared (Illumina Stranded Total RNA RiboZero Plus) with sequencing performed on the MGI DNBSEQ-T7 (∼50M reads, PE150bp). Longitudinal analyses in case samples (n=103) were conducted using the R package “variancePartition” with sex as a covariate. Gene expression changes associated with disease progression (ALSFRS-R) were modelled using regression analysis (matched with the subject and collection ID). DNA was extracted from blood collected in EDTA tubes and run on Illumina SNParray before standard quality control and imputation^39^. A large publicly available dataset AnswerALS data (v1, release5) was utilised for iPSC-MN transcript expression analyses^38^. Briefly, RNA was extracted (Qiagen RNeasy mini kit), libraries generated (Illumina TruSeq Stranded Total RNA library Ribo-Zero kit) and sequenced (Illumina Novaseq 6000, 50 million PE reads/sample). We analysed the raw read counts whereby transcripts had previously been aligned (GRCh38) and indexed (samtools) and quantified with featureCounts (subread v.2.0.1).

### Cross-Disease Comparison

We carried out a cross-disease comparison to compare genetic signals between the ALS chromosome 3 locus and other traits/diseases. To identify relevant traits/diseases the GWAS catalogue was searched for studies from 2010 onwards using the rsID (“rs631312”) and/or mapped genes (“*RPSA*” and “*MOBP*”). Eighty-one results were shortlisted, n=25 were independent studies and n=18 were independent traits (i.e. three were Amyotrophic Lateral Sclerosis and four were cortical thickness). This included brain imaging (n=11), neurodegenerative diseases (n=4) and others (n=3, bone mineral density, lean mass and liver pQTL). SuSiE fine-mapping was carried out on the neurodegenerative diseases with available summary data, this included ALS (as above), Frontotemporal dementia/FTD^15^ (N_cases_=4,685, N_controls_=15,308) and Corticobasal degeneration/CBD^16^ (N_cases_=219, N_controls_=3,750). We used LDlink to identify eQTLs of top SNPs across diseases that were in high LD (r^2^ > 0.8) in relevant tissues (brain, nerve, muscle and blood).

### Zebrafish maintenance

Zebrafish maintenance, crispant generation, observation, and phenotyping were conducted according to standard protocols approved by Griffith University (GRIDD1122AEC). Wild-type (AB), *Tg(MN:GFP)*^40^ and *Tg(kdrl:GFP ; gata1:dsRed)* lines - all on an AB background- were used. To model and benchmark motor neuron loss, we used maternal zygotic SMN-deficient mutants (*smnY262-/-*), which display early motor neuron degeneration as previously described^41,42^. In addition, a *TDP43*-deficient double-mutant (*tardbp-/-; tardbpl-/-*) which lasks both zebrafish orthologs of TDP-43 and exhibits progressive motor neuron pathology were developed in-house, mimicking the published double mutant^43^.

### Zebrafish Crispant Generation

CRISPR/Cas9 F0 embryos were produced as previously described^44,45^. Briefly, one-cell stage embryos were injected with a mixture of Cas9 protein (New England Biolabs, #M0646M) complexed with three different gRNAs. We used a 1:3 molar ratio (Cas9/gRNA) with an injected Cas9 dose of 800 pg final. Zebrafish *rpsa* CRISPR target sites and oligonucleotides used to generate CRISPR guide RNAs (**Supplementary Table 19**).

### Zebrafish morphology and motor neuron imaging

Injected animals were grown at 28°C and observed and imaged using an Olympus MVX10 mounted with a DP75 camera. To observe motor neuron development and axon pathology, on the day of analysis animals were dechorionated (as required), anaesthetised in tricaine and mounted in low-melting agarose in lateral view. Confocal images were taken using an Olympus FV3000 with images analysed in FIJI, with CaP-axon projection abnormalities scored. Stack snapshots were generated using Standard Deviation projection and results statistically compared using Welch’s t tests.

### Behavioural and Survival Experiments

Behavioural analysis was performed using a Zebrabox Revolution (Viewpoint) following the manufacturer’s instructions. Zebrafish larvae were distributed in 24-well plates filled with 500μL of E3 medium (one larva per well). Following the plate setup, all samples were placed at 28°C for a minimum of 1 h before running the analysis. The assays consisted of automatic recording of the larval swimming tracks/behaviour during 24 min and under alternating phases of 4 min light and 4 min darkness. At the end of the experiment, each larva was monitored to exclude potential dead animals from the data. Results were statistically compared using Mann-Whitney tests. Survival was monitored/recorded daily by manually observing each plate twice daily.

### Touch response assays

Touch-evoked response was induced by light mechanical stimuli/touch on the back of the larvae’s heads using metal forceps and scored following five repeats at 30-second intervals. At 30 hpf (hours post fertilisation), a coiling response was scored “1”, no response was scored “0”. At 48 hpf, the escape response was scored according to the total distance swum in response to the light touch: “0”, no response; “1” < 1 body length; “2” 1-3 body lengths; “3” >3 body lengths. Results were statistically compared using Mann-Whitney tests.

## Results

### Fine mapping methods show limited consensus

Three methods, FINEMAP, SuSiE and MsCAVIAR, were used to prioritise SNPs in each significant ALS GWAS region (**Supplementary Table 1, Supplementary Figure 1**). Credible sets in the *RPSA/MOBP* on chromosome 3 were identified in each method (**Figure 2**). FINEMAP (n=5 SNPs) and SuSiE (n=7 SNPs) had overlapping results (n=3 SNPs rs631312, rs1707981, and rs675595). MsCAVIAR identified an independent set of SNPs (n=5). With further analysis of the locus, the most associated GWAS SNP (rs631312) identified by MsCAVIAR and FINEMAP credible sets, varied based on the maximum number (n) of causal SNPs. For MsCAVIAR, rs631312 had a posterior PIP> 0.1 only when n was set to 1 or 4; when n was set to 2 or 3, the PIP for this SNP was close to 0. In contrast, FINEMAP included rs631312 in its credible set when n was set to 1 or 2. However, at n = 3, rs631312 was excluded. Notably, when n was increased to 4 or 5, rs631312 was again in the credible set with a PIP of 1. This observation underscores the sensitivity of fine-mapping tools to parameter settings and the importance of careful model specification.

**Figure 1:**
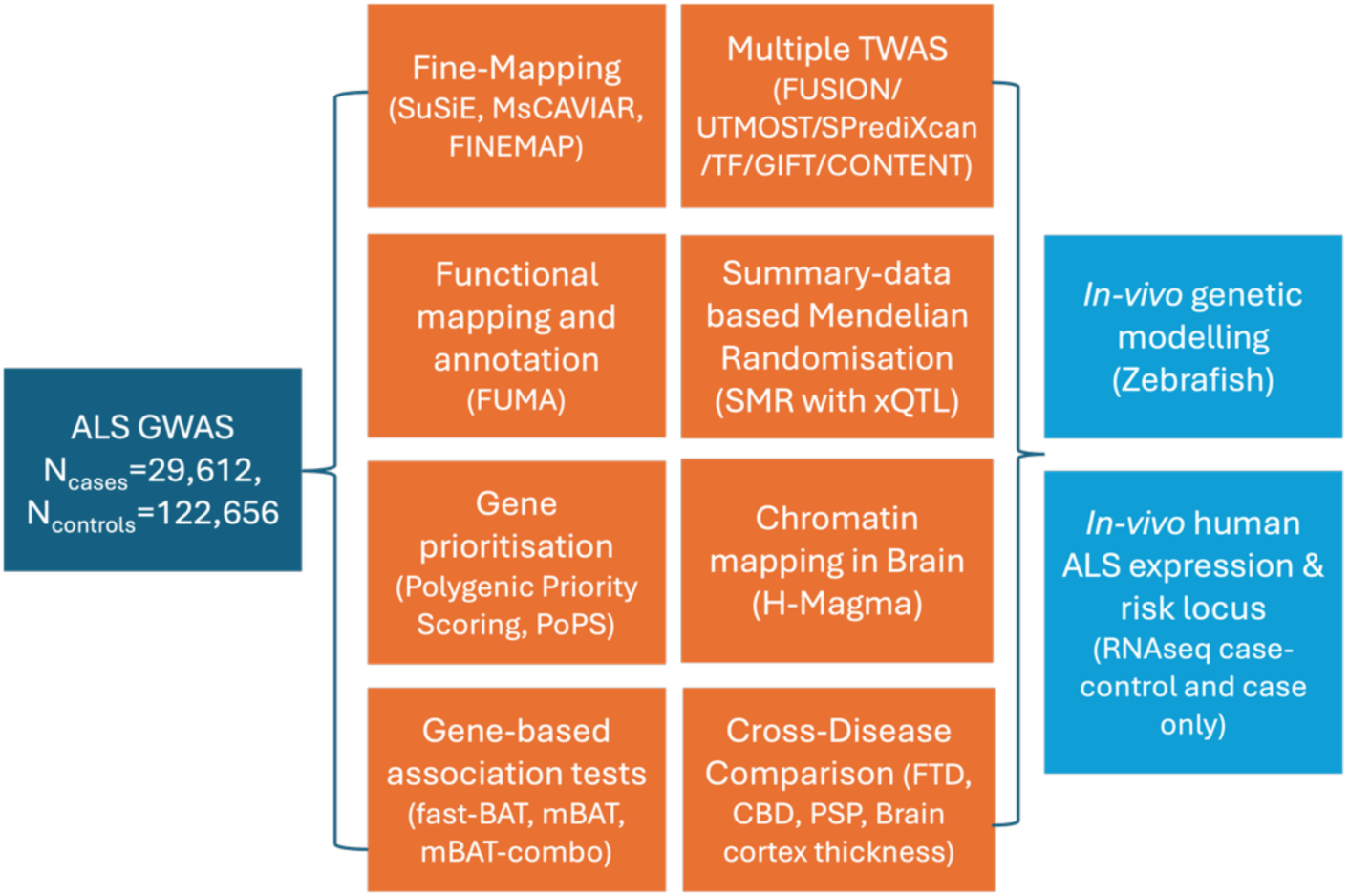
*In-silico* (orange) and *in-vivo* (blue) methods used to follow-up the *RPSA-MOBP* locus on the chromosome 3 identified in the ALS GWAS (dark blue). This included fine-mapping (estimating the posterior probability of causality for each variant and define credible sets likely to contain the true causal variant), Functional annotation and mapping (to annotate genes and biological pathways), gene prioritisation with polygenic priority scoring (integrating GWAS summary statistics with diverse functional genomic features to identify genes enriched for trait-relevant biological signatures), gene-based association testing (assessing the aggregated effect of the GWAS variants within each locus, to rank genes with the trait), transcriptome-wide association testing (integrating gene expression data to identify genes whose genetically regulated expression is associated with ALS;), summary- data based Mendelian Randomisation (using expression and splicing (xQTL) data to infer causal relationships, chromatin-informed mapping (leveraging chromatin accessibility and regulatory annotations in the brain (H- magma) to prioritise variants located in functionally active genomic regions) and cross-disease comparison (to compare genetic signals between ALS and other traits or diseases, to determine shared genetic mechanisms). These *in-silico* analyses were followed by analyses of ALS expression data and *in-vivo* validation using a Zebrafish

**Figure 2.**
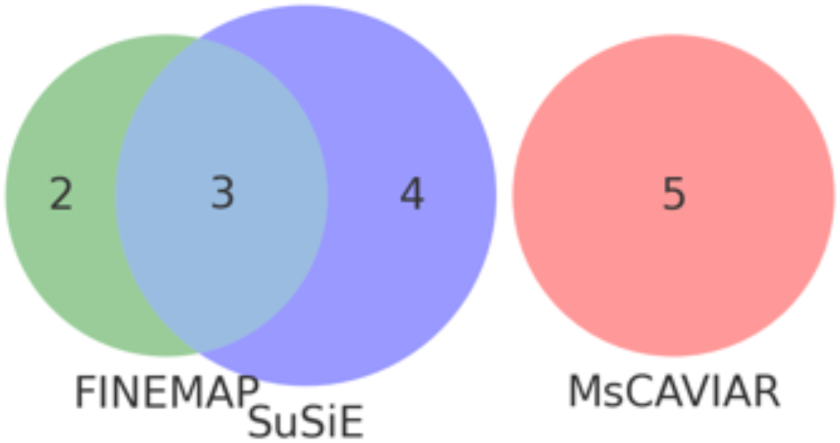
Fine-mapping methods generate distinct and overlapping results on Chromosome 3 (lead GWAS SNP rs631612). Visual of the SNPs identified by three methods in the chromosome 3 ALS GWAS. The size of the Venn diagrams represents the total number of prioritised SNPs identified, coloured by each method; FINEMAP (green), SuSiE (blue) MsCAVIAR (red). Despite using the same GWAS input, only FINEMAP and SuSiE shared prioritised SNPs (n=3).

### Functional annotation and positional mapping link four genes to the chromosome 3 locus

Both European-only and cross-ancestry ALS GWAS results were analysed in the FUMA pipeline (**Supplementary Table 2**). In the European GWAS (loci p < 5 x 10^-8^), FUMA identified 203 independent lead SNPs (**Supplementary Table 3**) across 61 genomic risk loci (p-value = 5 x 10^-6^). Using LD information (r² > 0.6), 2494 candidate SNPs were mapped for further annotation. Of the candidate SNPs, 61% were intronic, 20% intergenic with the remaining being a mix of exonic, non-coding, UTR or NA (**Supplementary Table 4**). 101 SNPs had CADD scores >12.37, suggesting potential deleteriousness. Over 65 SNPs were eQTLs in brain tissue datasets, including frontal cortex. On chromosome 3 locus, 14 genes were mapped, four with eQTL and chromatin data (*RPSA, MOBP, CXCR1, WDR48*) (**Supplementary Tables 2-5**).

Similar results were identified in the cross-ancestry GWAS. FUMA identified 215 independent lead SNPs (**Supplementary Table 3**) across 100 genomic risk loci (p-value = 5 x 10^-6^). Using LD information (r² > 0.6), 2757 candidate SNPs were mapped for further annotation. Of the candidate SNPs, 60% were intronic and 23% intergenic with the remaining being a mix of exonic, non-coding, UTR or NA (**Supplementary Table 4**). One-hundred and four SNPs had CADD scores >12.37, suggesting potential deleteriousness. On chromosome 3 locus, 15 genes were mapped, but as above in the European results, only four had both eQTL and chromatin data (*RPSA, MOBP, CXCR1, WDR48*) (**Figure 3**).

**Figure 3.**
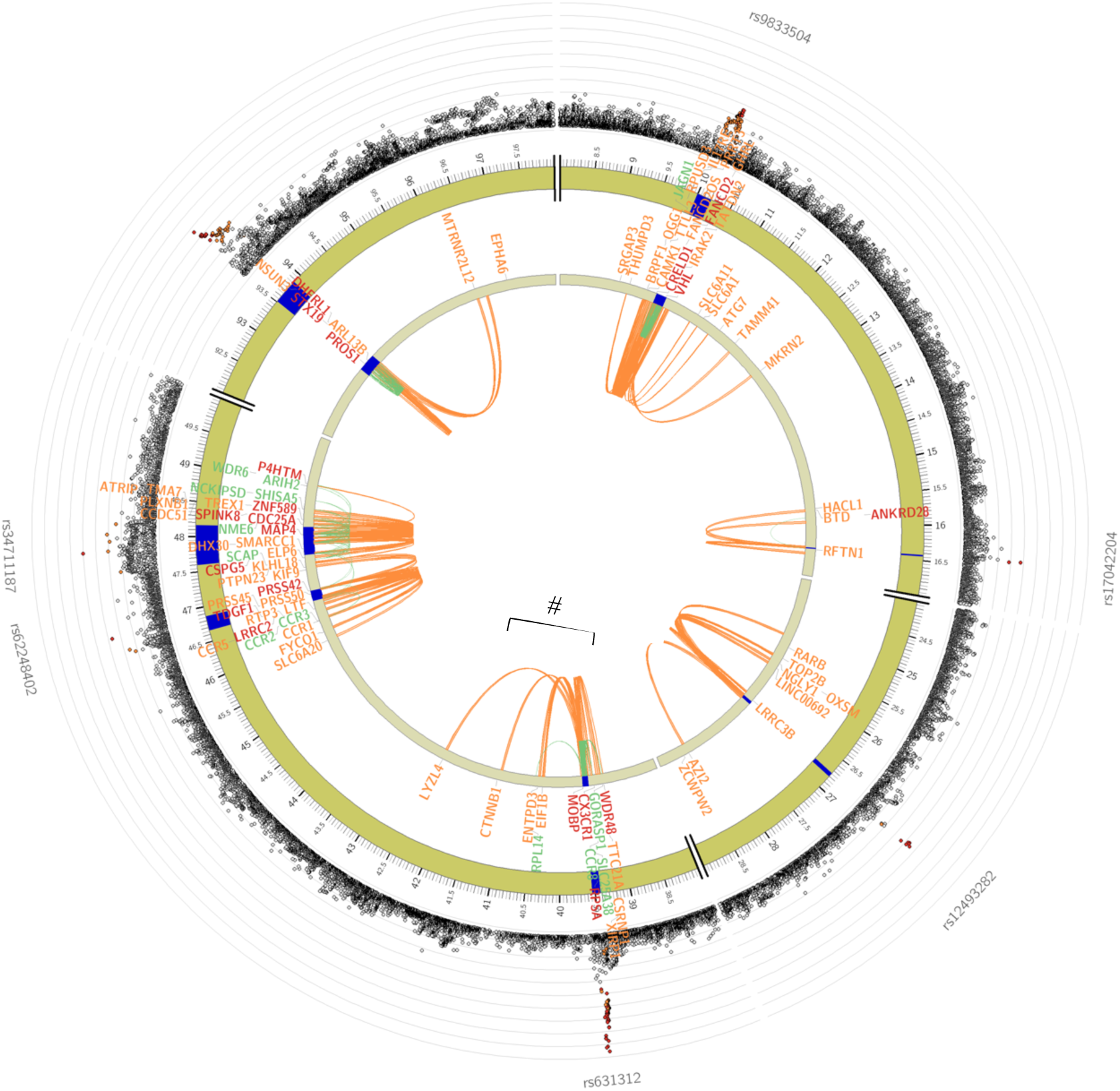
Circos plot of the Functional and Mapping annotation with cross-ancestry ALS GWAS summary statistics using FUMA. Highlighted with the Hash symbol and bracket is the region of interest. The outer dots indicate the log10 p-value with high LD SNPs with the lead SNP (rs631312) coloured red. Genes positionally linked with chromatin data are coloured orange, linked with eQTL data are green, and with both chromatin and eQTL are red (*MOBP, RPSA, WDR48, CX3CR1*). Linked details for each gene are in Supplementary Table 5.

### Gene-based association tests prioritised relevant gene on chromosome 3

Gene-based association tests (mBAT-combo^21^, MBAT and fastBAT^22^) identified 5 significant genes in the region (*MOBP, RPSA, SNORA62, SNORA63, SLC25A38*). MBAT-combo, given it is a combination of the other two tools, expectedly had the lowest p-values, with *MOBP* ranked 1^st^ (p= 9.31 x 10^-8^), *SNORA62* and *RPSA* ranked 2^nd^ and 3^rd^ (p=8.70 x 10^-6^ & 8.91 x 10^-6^ respectively) in the locus (**Supplementary Table 7**).

### Transcription-wide association studies identify multiple genes

For complementary insight we used a multitude of TWAS approaches. Across methods, we detected a total of 115 unique genes (**Supplementary Tables 8-10)**. First we ran TWAS-FUSION^23^ (FUSION framework, runs tissue-by-tissue), UTMOST (Unified Test for MOlecular SignaTures, models gene expression across multiple tissues), SPrediXcan (Summary-PrediXcan, run tissue-by-tissue but doesn’t account for correlation), TF-TWAS (Transcription Factor TWAS, includes transcription factor binding data to model gene regulation), and GIFT^27^ (Gene-level Inference of Functional Traits, approach that includes fine-mapping and uncertainty modelling). Fifty unique genes were significantly identified in these methods (significance threshold based on gene number was = 2.4×10^−6^). UTMOST had the largest number of signals, with its QQ-plot showing inflation/potentially picking up false-positives, that was not present using the other methods (**Supplementary Figure 24**). Only GIFT reported a signal in the chromosome 3 locus, finding *RPSA* as FDR significant (**Supplementary Table 8**). We further ran a conditional TWAS study, TWAS-CONTENT^28^. This approach can account for overlapping signals from nearby genes and correct for correlations between genes that are close together. Two strategies were used, one with CONTENT weights alone, and a second tissue-by-tissue approach. This detected 79 unique genes that passed Bayesian Bootstrapped FDR threshold (a hierarchical FDR to control within and across contexts) (**Supplementary Table 10**). None of the gene expression signals were strong enough to confidently link to the GWAS locus on chromosome 3. The genes *SLC25A38, RPSA and SNORA63* all had p-values <0.001, with *RPSA* and *SNORA63* identified in relevant tissues (brain and nerve) (**Supplementary Table 9**).

### Polygenic priority scoring (PoPS) ranks *RPSA* above other genes in the locus

Polygenic Priority Score^29^ was used to integrate signals from the ALS GWAS (gene-level association scores) together with ∼50K gene features (i.e. expression, pathway membership, protein interactions and regulatory elements) to provide a gene-based priority-score. The top 10 in each chromosome, as well as the chromosome 3 locus are reported using both the full GWAS cohort or EUR only (**Supplementary Table 12 and 13**). *RPSA* had a higher prioritised score than both *MOBP* and *SLC25A38* (*SNORA63* and *SNORA62* were not assessed). This suggests that *RPSA* shares more features with other GWAS hits, but did not feature in the top 10 chromosome results.

### Summary-data-based Mendelian Randomisation highlights *MOBP* but not *RPSA*

Genome-wide SMR (Summary-data-based Mendelian Randomization) analysis with xQTL data (based on methylation, protein and RNA expression/splicing) were carried out from the largest available blood and brain cohorts. This included eQTLgen (blood, N=31,684), BrainMeta (Brain, N=2,865, expression QTL, splice QTL), Interval, Fenland, SCALLOP (plasma protein QTLs n=10,708, n=3,301, and N=30,931) with methylation from BrainMeta (Brain, N=1,160) and Blood (LBC+BSGS, N=1,980). Using a genome-wide SMR threshold for each dataset and limited to those that pass the HEIDI test prioritised 9 candidates (*C9orf72, G2E3, MOBP, MYO19, RESP18, RPL26L1, SCFD1, SLC9A8* and *TPP1*) (**Supplementary Table 16, Supplementary Figures 14-23**). From the 12 loci detected, in the eQTL data there were 16 (out of 16) SMR significant genes, with 11 passing the HEIDI test. In the sQTL data, 14/16 SMR significant genes, with 9 passing the HEIDI test (**Table 1**). The smaller pQTL data, there was 1/16 SMR significant gene, which also passed the HEIDI test. In the mQTL data, 14/16 significant SMR genes with 12 passing HEIDI. This includes *MOBP* on chromosome 3, identified in the BrainMeta xQTL data (**Table 1**, **Figure 4**). Using a regional significance threshold (*p* value<0.0038), correcting for 13 genes within ∼1.5Mb of the GWAS locus of interest, *RPSA* is detected as a splice QTL in BrainMeta (**Figure 5**). However, this association does not pass the heterogeneity test (HEIDI, *P*=0.001) indicating that that the signal cannot be confidently distinguished between pleiotropy (a shared causal variant) and linkage (distinct causal variants in LD) (**Supplementary Table 16**).

**Table 1.**
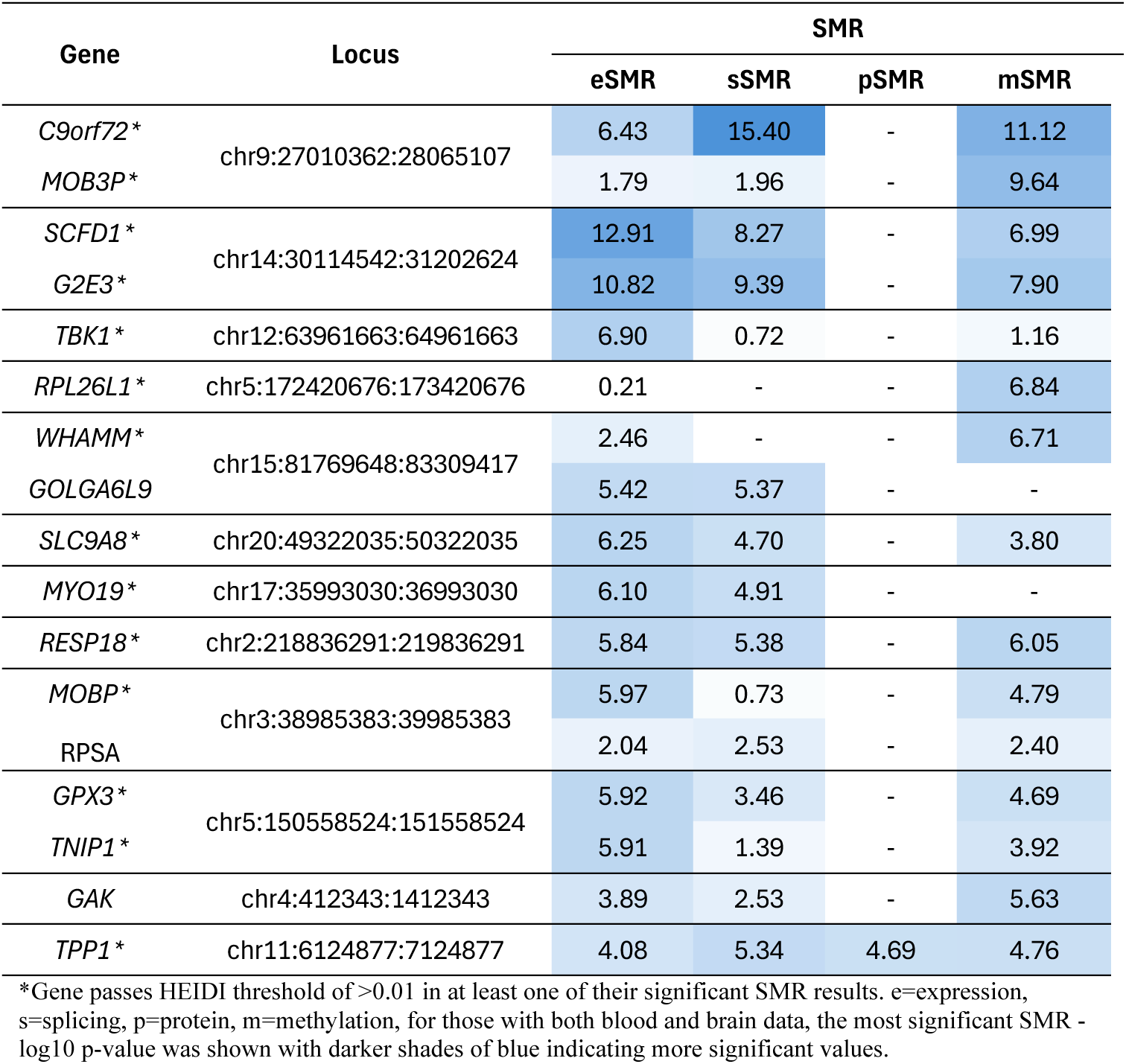

| Gene | Locus | SMR |  |  |  |
| --- | --- | --- | --- | --- | --- |
|  |  | eSMR | sSMR | pSMR | mSMR |
| *C9orf72* * | chr9:27010362:28065107 | 6.43 | 15.40 | - | 11.12 |
| *MOB3P* * |  | 1.79 | 1.96 | - | 9.64 |
| *SCFD1* * | chr14:30114542:31202624 | 12.91 | 8.27 | - | 6.99 |
| *G2E3* * |  | 10.82 | 9.39 | - | 7.90 |
| *TBK1* * | chr12:63961663:64961663 | 6.90 | 0.72 | - | 1.16 |
| *RPL26L1* * | chr5:172420676:173420676 | 0.21 | - | - | 6.84 |
| *WHAMM* * | chr15:81769648:83309417 | 2.46 | - | - | 6.71 |
| *GOLGA6L9* |  | 5.42 | 5.37 | - | - |
| *SLC9A8* * | chr20:49322035:50322035 | 6.25 | 4.70 | - | 3.80 |
| *MYO19* * | chr17:35993030:36993030 | 6.10 | 4.91 | - | - |
| *RESP18* * | chr2:218836291:219836291 | 5.84 | 5.38 | - | 6.05 |
| *MOBP* * | chr3:38985383:39985383 | 5.97 | 0.73 | - | 4.79 |
| *RPSA* |  | 2.04 | 2.53 | - | 2.40 |
| *GPX3* * | chr5:150558524:151558524 | 5.92 | 3.46 | - | 4.69 |
| *TNIP1* * |  | 5.91 | 1.39 | - | 3.92 |
| *GAK* | chr4:412343:1412343 | 3.89 | 2.53 | - | 5.63 |
| *TPP1* * | chr11:6124877:7124877 | 4.08 | 5.34 | 4.69 | 4.76 |

\*Gene passes HEIDI threshold of >0.01 in at least one of their significant SMR results. e=expression, s=splicing, p=protein, m=methylation, for those with both blood and brain data, the most significant SMR - log10 p-value was shown with darker shades of blue indicating more significant values.

**Figure 4.**
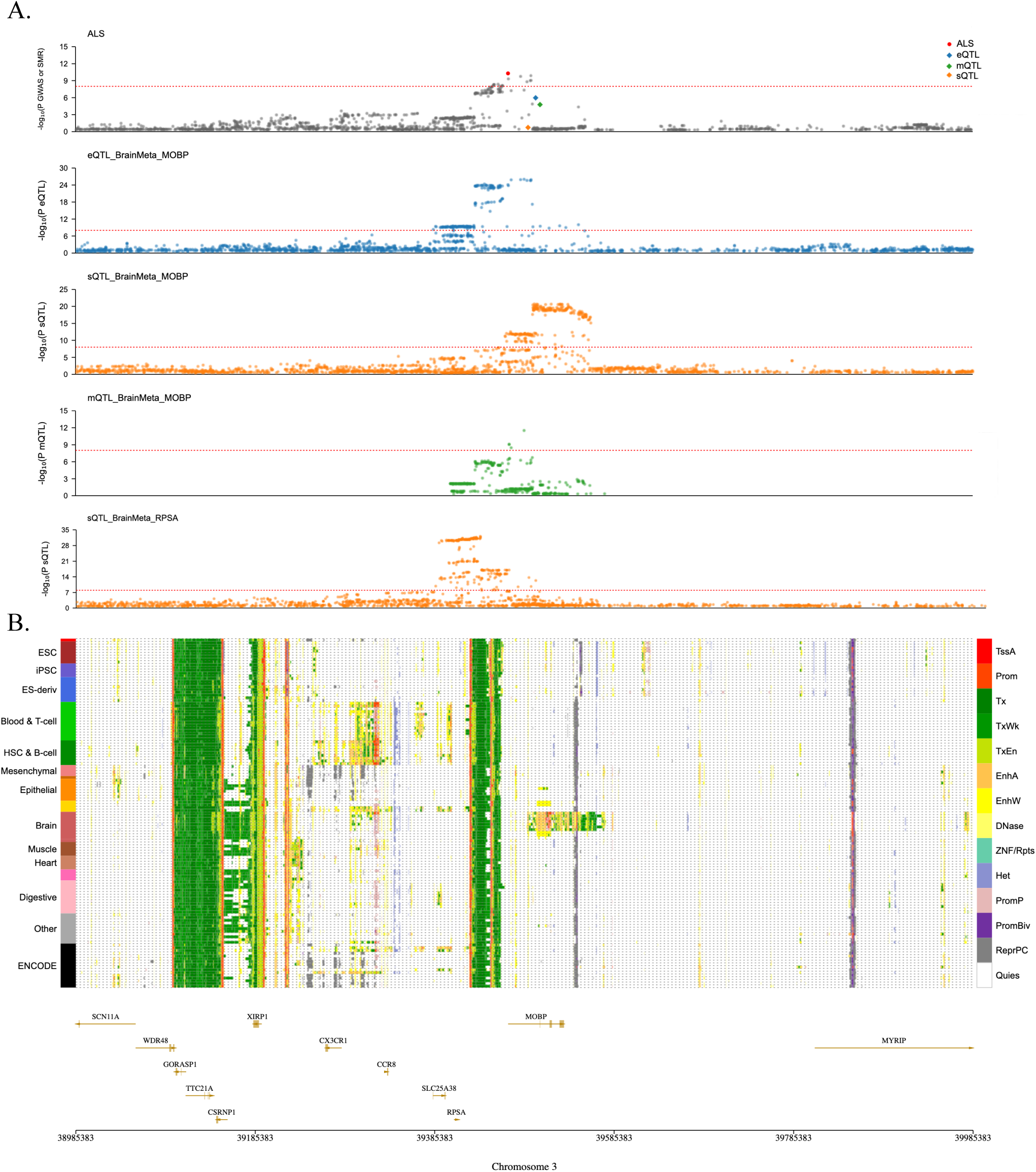
Chromosome 3 significant SMR results in brain detects *RPSA* and *MOBP* quantitative trait loci. A) ALS GWAS/SMR plot with *MOBP* results (passing HEIDI). Individual xQTL plots for significant SMR genes in the region. Plot of eQTL for *MOBP* (blue, -log10 *P* eSMR=5.97, BrainMeta, N=2,865), sQTL for *MOBP* (orange, -log10 *P* sSMR=0.73, BrainMeta), mQTL for *MOBP* (green, -log10 *P* sSMR=4.79, BrainMeta, N=1,160), sQTL for *RPSA* (orange, -log10 *P* sSMR=2.53, BrainMeta, N=2,865). The red dashed line indicates *p* value threshold of < 5 × 10⁻⁸. B) Epigenomic annotation of cells/tissues with corresponding gene map. A region overlapping the *MOBP* gene is specifically relevant to brain, while the intergenic region between *RPSA* and *MOBP*, and closest to the GWAS region, is ubiquitously regulated.

**Figure 5.**
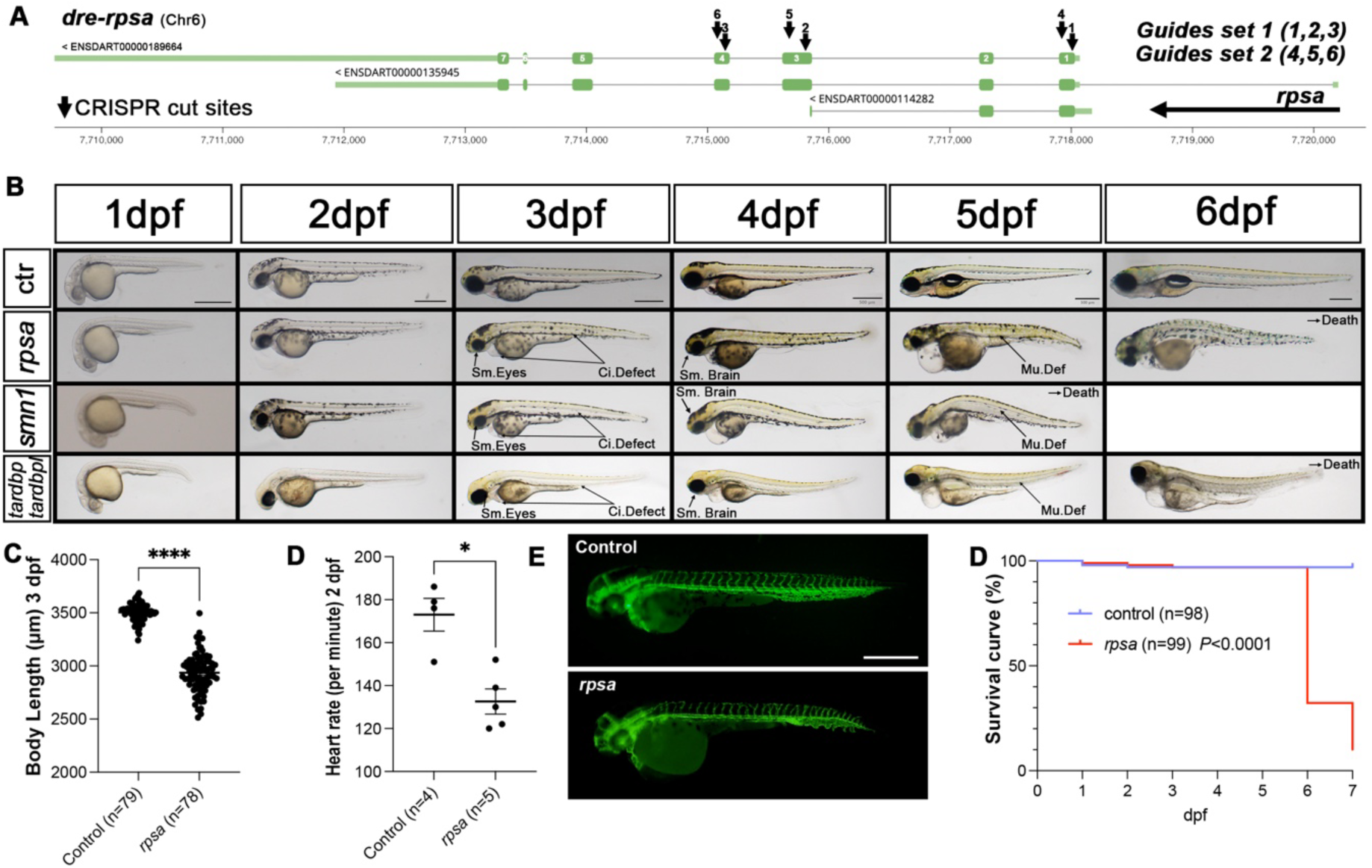
The *rpsa*-Loss of Function crispants mimics defects/regression observed in other MND models, the SMN- and TDP43-deficient zebrafish. **A)** The *dre-rpsa* genomic region and associated gRNA target locations. Two sets of guides were used, with set 1 combining guides 1, 2 and 3, and set 2 combining guides 4, 5 and 6 (**Supplementary Figure 26**). **B)** Snapshot of zebrafish larvae for Cas9 wild-type (wt) injected control (ctr), RPSA-deficient animals (*rpsa*), SMN-deficient (*smn1*) and TDP43-deficient animals (*tardbp/tardbpl* double mutants) Sm. Eyes= Small Eyes; Ci. Defect= Circulation Defects; Sm. Brain= Small Brain; Mu. Def= Muscle Defects. Scale bars, 500 µm. **C)** Body length of control versus RPSA-deficient animals at 3 dpf. **D)** Heart rate of control versus RPSA-deficient animals at 2 dpf. **E)** Snapshot of *Tg(kdrl:GFP)* animals evidencing control versus RPSA-deficient vasculature at 52 hpf. **F)** Kaplan-Meier survival curve of control versus RPSA-deficient animals.

### Chromatin-informed mapping of genes in Brain (H-MAGMA)

Using H-MAGMA, we identified gene-level associations with ALS GWAS using brain-specific Hi-C chromatin interaction data (**Supplementary Table 14)**. Significant associations observed in our locus of interest, included *RPSA* (FDR-adjusted p = 0.003) and *SLC25A38* (FDR-adjusted p = 0.006) (*MOBP* was not significant (p=0.11)) (**Supplementary Table 15**). These findings link relevant genes to distal non-coding variants through chromatin looping interactions, indicating a possible regulatory mechanism underlying their involvement in disease. Multiple SNPs were assigned to *RPSA* and *SLC25A38*, capturing long-range regulatory interactions otherwise missed by linear proximity-based methods.

### *In-vivo* human ALS expression data (RNAseq)

To examine the relationship between *RPSA* and *MOBP* expression levels in disease, we examined transcriptomic data in ALS case-control cohorts in blood (N=174 samples, N_cases_=121 cases, N_controls_=53). In blood, no difference in either *RPSA* or *MOBP* expression between ALS cases and controls was identified (*p*=0.64 and 0.39, respectively) was observed, noting that the mean count of *MOBP* was low across the cohort (average per sample 5.26) and not normally distributed. For samples that had matched genotype data (n=135), no associations were detected with lead risk SNP (rs631312, A=0.67, G=0.33) and *RPSA* or *MOBP* gene expression using a linear expression model (**Supplementary Figure 25**).

To examine changes over time for *RPSA* and *MOBP* in blood we examined longitudinal data in Australian ALS cases (n=41 individuals, n=103 observations, 2-4 visits). The change in ALSFRS-R in months since first visit and months since diagnosis we identified a linear decrease of 0.18 points (−0.31 to -0.06 95% CI, *p*=0.004) and 0.18 (−0.23 to -0.14 95% CI, *p*=3.51e-12) each month for *RPSA* and *MOBP* respectively (**Supplementary Figure 26**). Using a linear mixed-model analysis, fitting individual as a random effect, there was no association with expression and ALS functional rating score (ALSFRS-R) (scale from 48 to 0, where 48 is normal physical function) was identified (*p*=0.28) (**Supplementary Figure 27**).

We used an independent iPSC-MN cohort^38^ (n=255 samples, n=213 cases/n=42 controls) to examine *RPSA* and *MOBP* together with other known ALS Mendelian (*C9orf72, SOD1, TARDBP, TBK1*) and risk genes (*UNC13A, ATXN2*) in ALS cases and controls. Logistic regression was used to examine the association between expression and case-control status, adjusting for age and sex (**Supplementary Figure 28-29**). To test for the effect of expression and rate of progression, we limited the cohort to definite/probable ALS cases, who were of European ancestry, without a gene mutation and controlled for sex, age and onset (n=139). There was no change in the level of any known ALS gene with rate of progression (change in ALSFRS since onset) (p>0.05 **Supplementary Figures 28 and 30**).

### Cross-Disease Comparison

Genetic signals between the ALS chromosome 3 locus and other traits/diseases were identified using the GWAS catalogue (n=25 studies/18 independent traits) (**Supplementary Table 22**).

We further analysed four neurodegenerative diseases (n=4) given expected shared signals. Each top SNP was either the same hit or had strong linkage (D′ > 0.93) with the top ALS SNP (rs631312). The correlation between each allele was high (r^2^>0.98) except the FTD SNP which was moderate (r^2^=0.31) (**Supplementary Table 18**). SuSiE fine-mapping was carried out on three with available summary data, including ALS (as above), Frontotemporal dementia/FTD^15^ (N_cases_=4,685, N_controls_=15,308) and Corticobasal degeneration/CBD^16^ (N_cases_=219, N_controls_=3,750). CBD and ALS shared credible SNPs but not ALS and FTD (**Supplementary Table 18**). The GWAS signals for the neurodegenerative diseases and cortical thickness colocalise with eQTLs in tibial nerve, skeletal muscle and brain with strong effect sizes and very low p-values. The most significant was in the tibial nerve, top eQTL SNP for *MOBP* was rs616147, B= -0.8, p-value=2×10^-56^, and *RPSA* was rs528364, B= 0.4, p-value=2×10^-34^. In skeletal muscle, top eQTL SNP were rs528364, B= 0.2 *RPSA*, p-value=2×10^-18^ and in brain-cortex (top eQTL SNPs were in high LD with rs674192/rs675595, B= 0.2 *RPSA*, p-value=1×10^-^ _5_) (there was no eQTL signal for *MOBP* in muscle or brain).

### *rpsa*-Loss of Function zebrafish mimics pathology observed in SMN- and TDP43- deficient zebrafish

To investigate the impact of *RPSA in vivo*, we deployed a robust bi-allelic knockout approach recently developed for the zebrafish animal model^44–46^. Interestingly, the *RPSA* homolog in the zebrafish (*dre-rpsa* or *rpsa*) encodes a protein with a striking >90% homology with its human protein counterpart, suggesting strong conservation (**Supplementary Figure 31**). We used Cas9 protein injections complexed with three guide RNAs targeting *rpsa* exons 1, 3 and 4 (**Figure 5A**). To ensure robustness of our approach, we conducted independent injections with two sets of guides, with both sets leading to the same results as presented below (**Figure 5A**, **Supplementary Table 19**). While *rpsa*-crispant zebrafish develop normally in the very early stage, the animals develop striking abnormal morphological features phenocopying defects/pathology observed in other zebrafish models of motor neuron diseases, *e.g.* SMN- and TDP-43-deficient animals (**Figure 5B**)^47,41,42,43^. From 3 dpf (days post fertilisation), animals present with smaller eyes, circulation defects (average heart rate *rpsa*-crispant 132.6 ±5.9 vs control 173 ±7.6 bpm (beats per minute), P<0.05) and shorter length (*rpsa*-crispant 2,936 ±20.89 vs control 3,503 ±9.41µm, P<0.0001) (**Figure 5C, D and E**). From 4 dpf, animals develop cardiac oedema and muscle fibre abnormalities. These features appear to develop with similar chronology and severity in RPSA, SMN and TDP-43 deficient animals. TDP-43 deficient larvae have, in addition, an apparent absence of pigmentation that is not observed in the two other conditions (**Figure 5C**)^43^. In terms of survival, *rpsa*-crispant animals develop pathology that culminates in death between 6/7 dpf, slightly longer than SMN-deficient larvae (5/6 dpf) but slightly shorter than TDP43-deficient animals (7/8 dpf) (**Figure 5E**)^41–43^.

### *rpsa-*LoF triggers axon pathology and progressive loss of motor function

To further explore the pathogenicity triggered by *RPSA*-deficiency in zebrafish, we analysed motor neuron anatomy as well as motor function in our zebrafish model. As presented in the **figure 6A** snapshots, *rpsa*-LOF triggers obvious morphological changes of the zebrafish caudal primary (CaP) motor neuron as early as 30 hpf. A deepened observation of CaPs 7 to 12 (**Figure 6A**) demonstrated a significant reduction of their axon length with an average length of 98.18 (±1.58) µm for *rpsa*-LOF samples versus 87.02 (±2.27) µm for control (**Figure 6B**). Similarly, CaP axons in *rpsa*-crispants are significantly abnormally branched, a typical axonal pathology phenotype also triggered by SMN and TDP43-deficiency in zebrafish (**Figure 6C**)^41–43,47^. While animals develop early hallmarks of axonal pathology, they do respond normally to touch with no difference to control at both 1 and 2 dpf (**Figure 6D**), suggesting a normal wiring of the brain as well as an overall functional neuromuscular system. Although *rpsa*-crispant animals present normal motor function patterns up to 3 dpf (**Figure 6E and F**), they develop rapid and progressive motor decline from 4 dpf onward, which culminates with premature death before 7 dpf as previously described (**Figure 6E and F**). Across a 24 minute recording in 24-well plate, with loops of 4 minutes of light and 4 minutes of darkness (stimulating swimming activity), *rpsa*-crispants did not significantly differ from controls at 3 dpf with a distance swum of 7.9 (±2.33) mm vs 6.3 (±1.93) mm for the controls, which rapidly declined up to death (**Figure 6E and F**). The presence of early axonal pathology combined with progressive loss of motor function that culminates with early death strongly mimics previously published models associated with disorders of the motor neurons, strongly supporting *RPSA* as an essential gene in motor neuron biology and a plausible key modulator of ALS pathophysiology.

**Figure 6.**
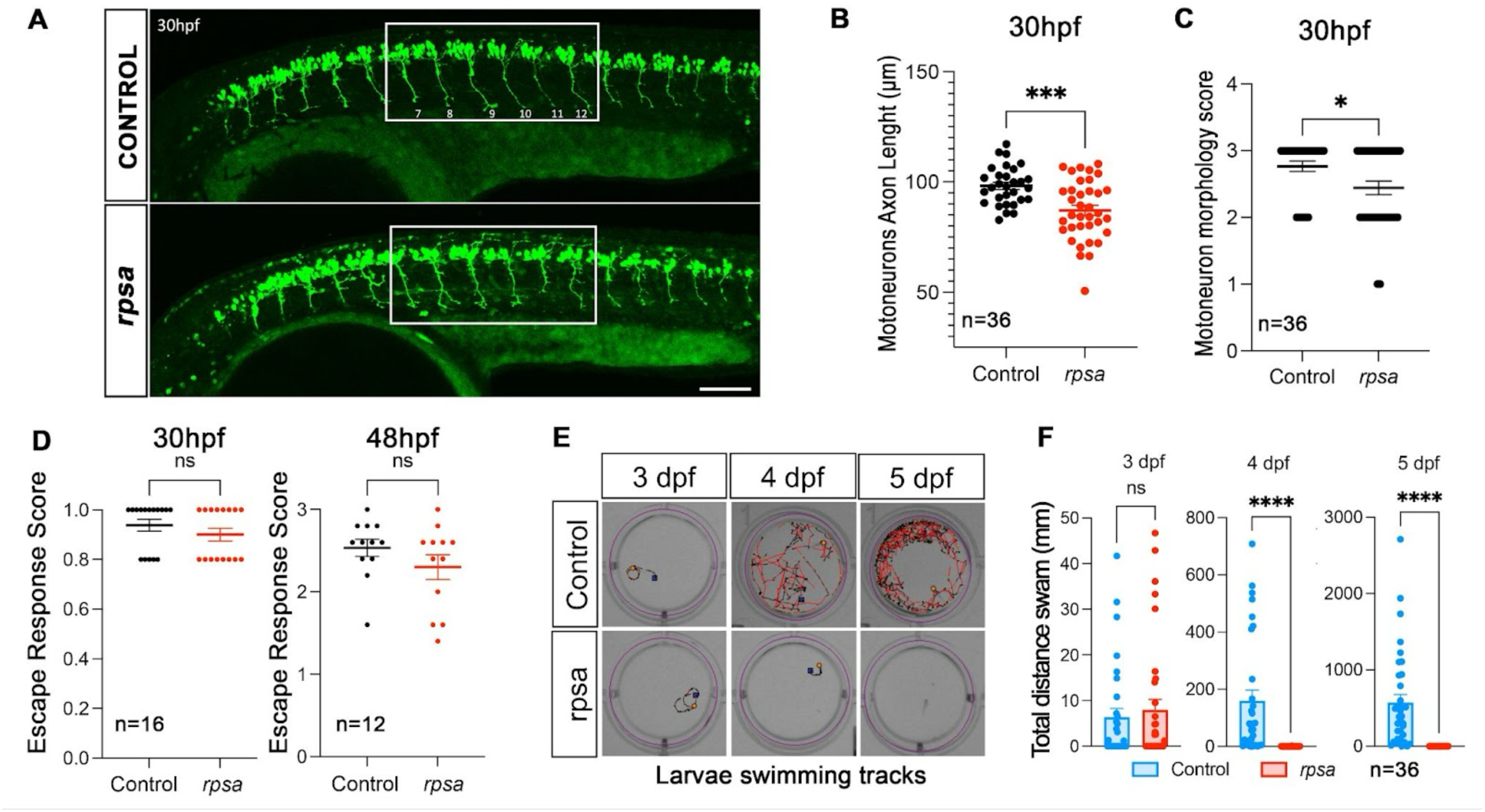
The *rpsa*-knockout triggers motor neuron axon pathology and progressive loss of motor function. **A)** Confocal standard-deviation projection of control versus RPSA-deficient animals highlighting the motor neuron axonal pathology triggered by *rpsa*-LOF. Caudal primary (CaP) axons 7 to 12, as labelled, were further analysed to quantify the observed obvious axonal defects. Transgenic *Tg(MN:GFP)* line expressing eGFP in motor neurons. **B)** CaP motor neuron axon length in control versus RPSA-deficient animals demonstrates significantly shorter size. **C)** Compared to control, CaP motor neuron axons in RPSA-deficient animals are abnormally branched, a typical feature of axon pathology observed in SMN- and TDP43-deficient animals. **D)** Touch-evoked escape assays at 1 and 2 dpf. RPSA-deficient animals respond normally to touch at 1 and 2 dpf, suggesting a normal early development and functional neuromuscular system and brain function. **E)** Snapshot of swimming tracks/patterns of control versus RPSA-deficient animals at 3, 4 and 5 dpf through a 24-minute recording with alternating 4 minutes of light and 4 minutes of darkness. **F)** Swimming track/distance quantification of the video recording demonstrates no motor function difference at 3 dpf followed by a rapid and progressive decline of motor function at 4 and 5 dpf in *rpsa*-deficient animals.

## Discussion

ALS is a genetically and clinically heterogenous neurodegenerative disease with both known and unknown risk loci contributing to disease. For complex conditions, genome-wide association studies have been instrumental in uncovering risk variants and nominating causal regions that may inform future drug-discovery^7,8,12^. Here, we focus on the chromosome 3 *RPSA-MOBP* locus (lead SNP: rs631312, OR=1.083, 95% CI: 1.06–1.10), integrating comprehensive *in silico* and *in-vivo* functional data to prioritise risk genes. The genomic analyses were not restricted to this region and that many of the *in-silico* analyses detected relevant/known ALS genes (**Supplementary Table 17**) to support this approach. Our findings for chromosome 3 converge on *RPSA* as a plausible contributor to ALS pathogenesis, warranting further investigation into its biological role and potential relevance to therapeutic development.

Fine mapping of the ALS GWAS locus on chromosome 3 revealed that the lead SNP, along with two others, were consistently identified by two of the three methods used (FINEMAP and SuSiE). However, sensitivity analyses—where the maximum number of causal SNPs at the locus was varied i.e. from one to four—showed that the posterior inclusion probability (PIP) for the lead SNP fluctuated between 0 and 1. This variability highlights the strong influence of parameter settings and model specifications on fine-mapping results, making it difficult to draw definitive conclusions. The limitations of current methods, particularly their inability to adequately account for cohort heterogeneity (typical in large multi-national consortia) have been underscored in a recent meta-analysis applying multiple fine-mapping approaches^48^.

Given the absence of a single method considered definitely accurate for determining likely causal genes with high accuracy, we used a variety of complementary methods to prioritise relevant genes in the region. To briefly summarise, functional annotation of the ALS GWAS summary statistics (FUMA) identified four genes linked via expression and chromatin data: *RPSA, MOBP, CX3CR1, WDR48.* Gene-based association testing similarly detected *MOBP (p-value: 1.66E-07), RPSA (9.35E-07), SLC25A38 (1.59E-05)* without prioritising *CX3CR1 (0.08).* TWAS analyses for the region were relatively unpowered with limited findings, only GIFT highlighting *RPSA.* Polygenic priority scoring had the highest scores for *RPSA* and *CX3CR1* (=>0.2), with others (*SLC25A38*, *MOBP*, *WDR48*) close or below zero*. H-MAGMA* (using chromatin data from Brain tissue) prioritised *RPSA* and *SLC25A38*. Integrating expression, splicing and methylation data in a formal statistical framework (SMR) highlighted *MOBP* in brain tissue. This association also passed the HEIDI test, consistent with a single causal variant model rather than distinct signals in linkage disequilibrium (LD). However, we note that while HEIDI reduces the likelihood of LD confounding, it does not definitively identify the causal SNP- the top ranked SNP may simply tag the causal variant through LD. *RPSA* was also linked in SMR but did pass the HEIDI threshold (meaning LD-driven association cannot be excluded). Notably, prior SMR analyses reported similar findings^8^ but lacked integration of splicing QTLs or HEIDI-based heterogeneity testing. Collectively, our results consistently prioritise *MOBP* and *RPSA* across multiple *in-silico* methods, warranting functional follow-up.

Additional interrogation of the lead ALS SNP (rs631312) and its surrounding LD structure further supports potential regulatory activity in disease-relevant tissues. Notably, rs631312 is in moderate to high LD with several functionally annotated variants, including rs1768190, which has been identified as a strong eQTL for both *MOBP* and *RPSA* in the tibial nerve (GTEx v8). While *MOBP* exhibits a highly brain- and spinal cord-specific expression profile, *RPSA* is more ubiquitously expressed, with strong eQTL effects across multiple tissues, including tibial nerve (min p = 7.6×10⁻¹⁰⁰) and skeletal muscle, where it also shows significant sQTLs (min p = 5.0×10⁻¹⁷⁸). Interestingly, the shared regulatory activity in the tibial nerve aligns with known early ALS pathology in peripheral neurons, raising the possibility that genetic risk at this locus could influence both central and peripheral neurobiology.

These findings suggest that regulatory variants in the region may modulate both RPSA and MOBP expression in tissue-specific contexts, potentially contributing to disease via multiple mechanisms. The functional overlap in tibial nerve expression also raises the hypothesis that a single causal variant could exert pleiotropic regulatory effects, which may explain why both genes are recurrently prioritised across different analytic platforms. Future studies using fine-mapped eQTL data in disease-relevant tissues, particularly spinal cord or peripheral nerves, will be important to further dissect these regulatory mechanisms.

*MOBP* encodes the myelin-associated oligodendrocyte basic protein gene, with a highly specific central nervous system expression pattern, where it is limited to the brain and spinal cord. It is abundant in the central nervous system of mammals and as such, *mobp-*deficient mice have previously been created to understand its role^49,50^. Phenotypic analyses of a knock- out (KO)-*mobp* mouse model^50,51^ showed no significant effects (*i.e.* normal overt mouse) with the exception of changes in axon diameter. An independently generated KO-*mobp* model^49^ did not replicate this finding, and showed unchanged compact myelin^49^, unperturbed myelination, no deficit in oligodendrocyte number and myelin sheath thickness^49^. Under stress (exposure to a toxic myelin substance (HCP)) may impact KO mice more (imaging found more split myelin sheaths compared to WT) which might be due to altered arrangement of the radial component^51^ but this has not been replicated. Current literature suggests that subtle changes in *mobp* expression may have very limited/if any effects on CNS function, as only when KO/under stress has a phenotype been recorded.

*RPSA,* which encodes a ribosomal protein SA, is a multifunctional protein with ubiquitous expression across tissues/cells. In addition to its role as a 40S ribosomal subunit, RPSA also functions as a laminin receptor, mediating processes such as cell adhesion, differentiation, migration and neurite outgrowth, as well as RNA binding and ribosome assembly. While not previously linked to ALS, *RPSA* has potential disease relevance through interactions with key ALS proteins such as TARDBP^52^ and SOD1^53^. With recent relevance to Alzheimer’s disease pathology^54^, converging evidence exists for a potential role in neurodegeneration.

Highly conserved across species, including zebrafish (orthologue with >90% homology), supported a functional follow-up investigation in this versatile organism. Our study shows that CRISPR/Cas9-induced *rpsa-*LoF in zebrafish triggered progressive and severe phenotypes which mimicked the pathology observed in well-characterised zebrafish lines associated with key motor neuron disease proteins (SMN and TDP43). Specifically, in the *rpsa*-ko, we found marked motor neuron axon pathology, progressive loss of motor function and rapid decline culminating with premature death prior to 7 dpf. These also presented with prominent cardiovascular abnormalities, defects also observed in both SMN- and TDP43-deficient models^47,41,42,43^. These *in vivo* results strongly support *RPSA* to be a likely contributor to ALS pathophysiology.

There were no clear transcript expression changes in blood in preliminary analyses of ALS cases and controls. To improve sensitivity to disease-relevant signals, we next examined an iPSC-MN^38^ dataset; however, this analysis similarly did not identify disease specific changes. We also tested other known ALS-associated genes, but none showed expression changes in this context. These findings suggest that transcript-level alterations—at least in the tissues and sample sizes examined—may not be a major driver of disease, or that their effects may be too subtle to detect in our current datasets. Given that the lead GWAS risk variant is a known eQTL for *RPSA* and *MOBP*, future work in larger, tissue-specific cohorts (e.g. spinal cord or single-cell contexts) may be required to more definitively assess expression-based mechanisms.

In meta-analysed iPSC-derived motor neurons from ALS and controls, *RPSA* had the highest mean levels of expression, compared to other risk genes and known ALS genes. No fold changes were detected between ALS vs. controls, but this was also not detected in other risk/causal ALS genes (**Supplementary Table 20**). In post-mortem spinal cord, many known ALS genes were significantly changed. In this dataset, *MOBP* had the highest mean expression and was decreased in ALS. This is consistent with loss of motor neurons and myelin, where *MOBP* is specifically expressed.

Our work has a number of limitations. The most relevant (given almost all our methods rely on this) is the ALS GWAS. Despite being the largest cohort available (N>130K) the ALS risk signal is still emerging. The SNP-based heritability is relatively modest with estimates such as h_obs_^2^ = 0.028, s.e. = 0.003 or h ^2^=0.043 (s.e. ∼ 0.0046), assuming a population lifetime prevalence of 0.25%. These estimates reflect the heritability captured by common variants rather than the total genetic contribution to ALS. We expect that larger meta-analysed datasets will help to refine these signals further. Specific for the *RPSA/MOBP* locus on chromosome 3, where we apply multiple *in-silico* tools to the region, our functional studies did not model all genes in the region, instead focussing on those consistently prioritised. We were unable to model ko-*mobp* in zebrafish (no orthologue was identified) and rely on previous literature of its function (two independently generated KO-*mobp* mouse lines). Our *in-silico* analyses did not integrate single-cell (SC) data but recent novel methods utilising the ALS GWAS^55^ suggests that a stronger ALS GWAS signal is necessary to see cell-specific results in a spatiotemporal context. Similarly, we note the cohorts with tissues relevant for ALS pathology (i.e. brain) remain small compared to blood, which may limit findings.

As yet, there remains no single method that can identify the most relevant risk genes in a particular disease. The pipeline and *in-silico* analyses here have been carried out in a genome-wide and in a complementary manner to support follow-up of *RPSA/MOBP* locus and other loci. The findings across loci serve an important purpose, sensitivity to benchmark tools i.e. when looking at a known gene/in a risk locus such as *C9orf72* and also potentially highlighting other genes, that might not have been previously considered by the field (summarised in **Supplementary Table 17)**. Previous ALS studies have carried out some of these analyses^8,56^ but using different datasets and approaches (i.e. SuSiE/TWAS-FUSION)^56^. Our work extends these to include additional methods and complementary analyses adding novel findings (i.e. the combined TWASs detect 30 overlapping, 85 distinct genes from previous report)^56^ and we generate and phenotype a functional model.

Our focus on the *RPSA/MOBP* locus was of particular interest because of the shared genetic risk loci across corticobasal degeneration (CBD), progressive supranuclear palsy (PSP), and frontotemporal dementia (FTD). Fine–mapping available data demonstrated credible set overlap between CBD and ALS, but not between FTD and ALS- potentially due to methodological limitations and/or study power. When we examined the strongest GWAS signals in each neurodegenerative disease (in some cases identical to ALS) showed strong linkage disequilibrium and large cis-eQTLs in tibial nerve (*MOBP/RPSA*), muscle (*RPSA*) and brain (*RPSA*).

In PSP, *MOBP* has been nominated as the causal gene in the locus^17^ based on evidence from oligodendrocyte single cell data and downregulation in PSP brains, in line with its role in synthesis and maintenance of myelin. However, LoF models do not support a major role for this protein in myelin function, raising questions about its importance in myelin integrity.

The shared associations across a number of traits/diseases highlights the roles of pleiotropy (a single locus influences multiple phenotypes) and diagnostic heterogeneity in neurodegenerative disease (since overlapping clinical and pathological features complicate case definitions in genetic studies). Pleiotropic effects have been linked with other loci such as *C9orf72, MAPT,* and *TMEM106B.* Shared genetic risk loci may represent core neurodegenerative mechanisms that transcend traditional disease classifications, warranting further investigation into molecular pathways underlying these links.

## Data availability

Any data produced in the present study are contained in the manuscript, available online or with relevant approval. The summary statistics used for the ALS GWAS were sourced from NHGRI-EBI GWAS catalogue at https://www.ebi.ac.uk/gwas/ (accession IDs GCST90027163 and GCST90027164 for cross-ancestry and European ancestry meta-analyses, respectively). The Frontotemporal dementia GWAS was downloaded from the UCL Research Data Repository (https://doi.org/10.5522/04/25600692.v1), the Corticobasal degeneration/CBD GWAS was provided following a direct request to corresponding author. At the time of submission, the RNA-seq blood data are part of a separate manuscript (in preparation) and will be made publicly available upon its publication.

## Funding/Acknowledgements

This project and the data generated was funded by a Daniel McLoone MND Research Grant from Motor Neurone Disease Australia (IG2312, to FCG). Additional project funding and data support were provided by an IMPACT grant from FightMND (2022, to FCG) and by the (Australian) National Health and Medical Research Council grants (grants 1078901, 1113400, 1087889, to NRW). MJT was funded by an EMBO Fellowship (ALTF 266-2023), JG; an NHMRC Investigator grant (APP1174145) and FCG; a Scott Sullivan Fellowship (MND and Me/MNDRA). We would like to acknowledge authors that shared the CBD summary data (Naomi Kouri, Owen A. Ross, Dennis W. Dickson and Gerard Schellenberg).

## Conflicts of interest

None declared.

The authors confirm that there are no financial or non-financial interests or relationships— such as consultancy, employment, honoraria, patents, personal relationships, stock ownership, or funding from commercial entities—that could be perceived to influence the objectivity or integrity of the research findings.

## Notes

### Competing Interest Statement

The authors have declared no competing interest.

### Author Declarations

Metro North Health Human Research Ethics Committee EC00172 of the Royal Brisbane and Women's hospital gave ethical approval (2006/047) for this work. The University of Queensland Human Research Ethics Committee gave ethical approval for this work (2021/HE002682).

