## Supplementary Figures for "Genomic and Functional studies identify *RPSA* as a risk gene for ALS and other neurological diseases"

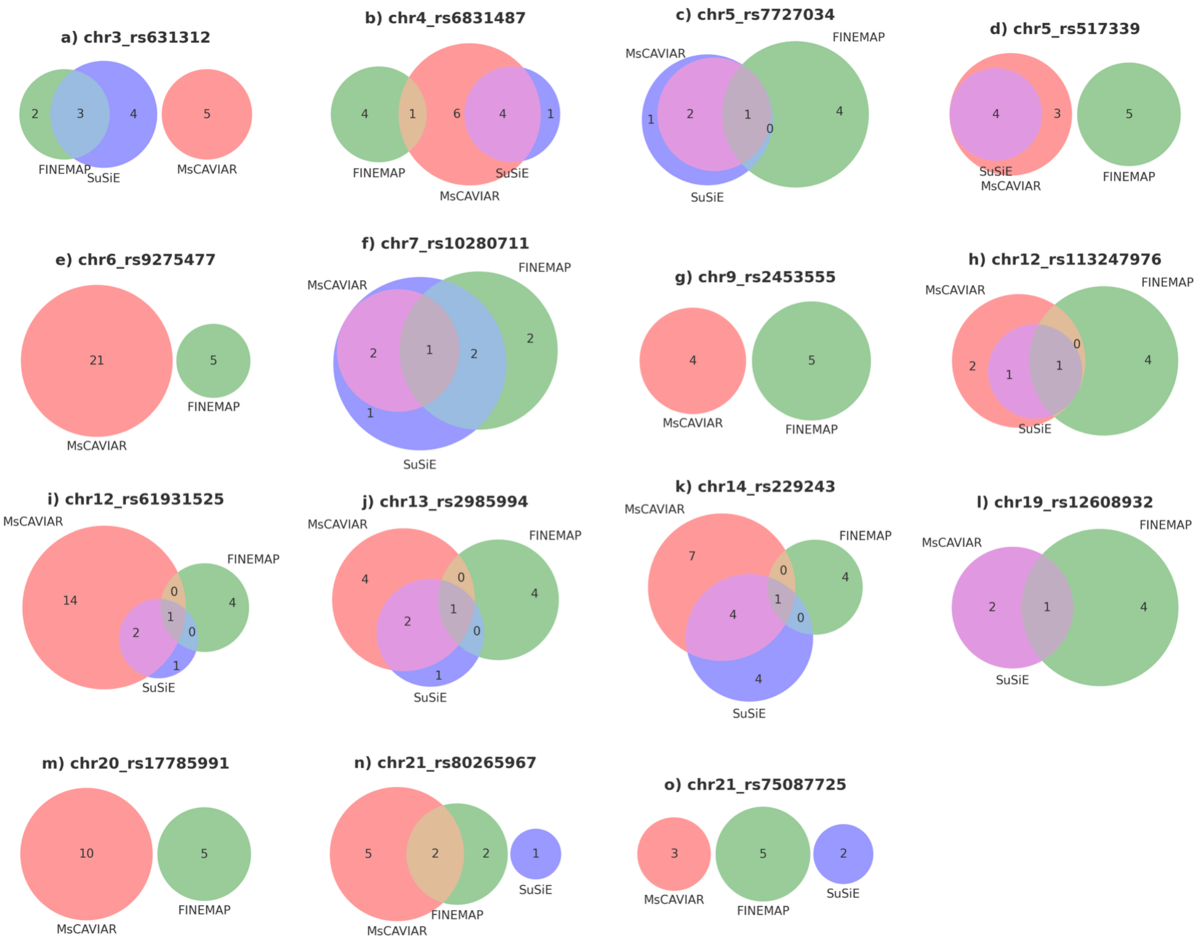


**Supplementary Figure 1: Overlap of SNPs identified by three fine mapping methods.** Venn diagrams illustrate the shared and unique SNPs identified by MsCAVIAR (red), FINEMAP (green), and SuSiE (blue) in 15 genomic regions (panels a-o). The lead SNP for each region is denoted in the title of each panel. Numbers within the Venn diagrams represent the count of SNPs uniquely or commonly identified by the methods.

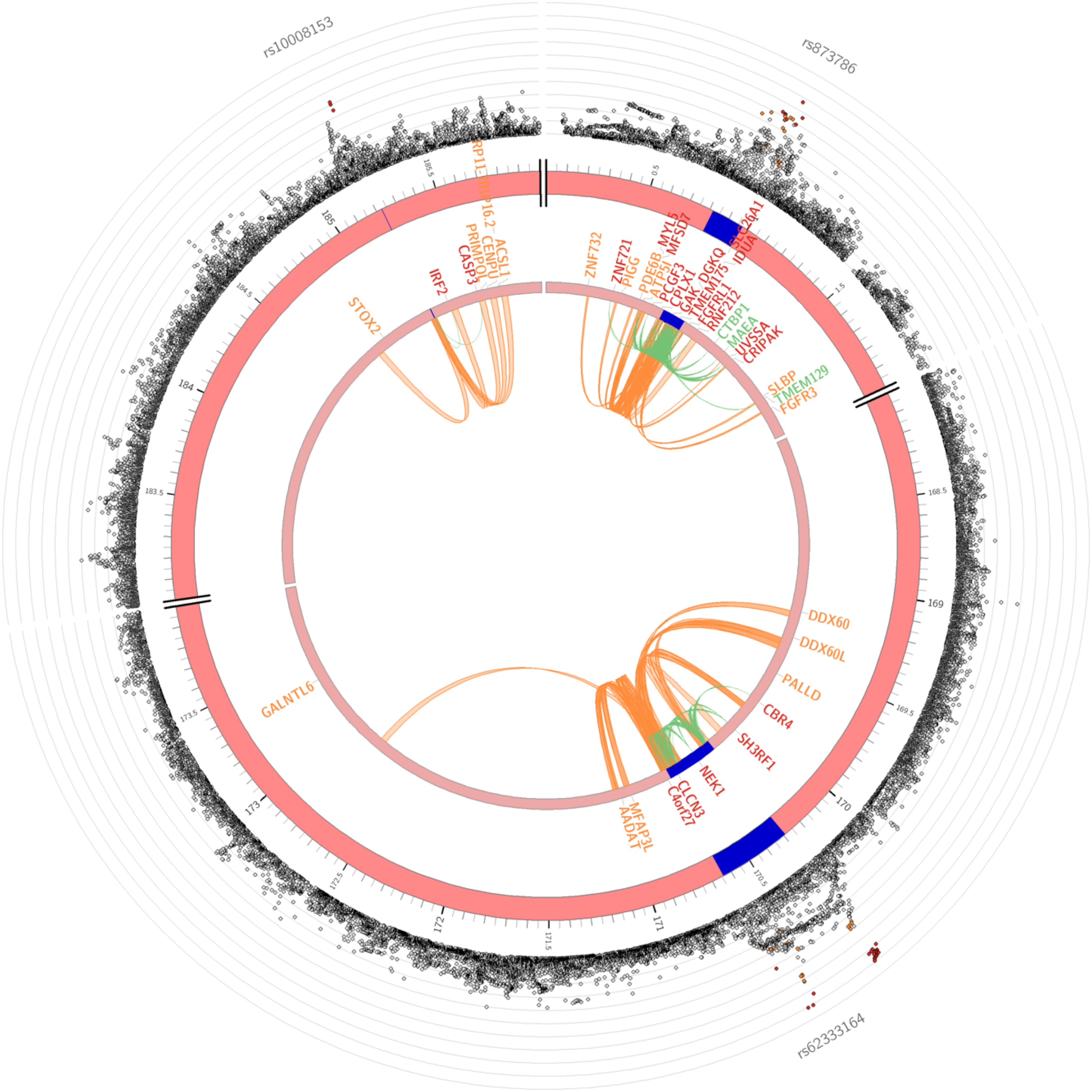


**Supplementary Figure 2**. **Circos plot of the Functional and Mapping annotation with cross-ancestry ALS GWAS summary statistics using FUMA on chromosome 4.** The outer dots indicate the log10 p-value with high LD SNPs with the lead SNP coloured red. Genes positionally linked with chromatin data are coloured orange, linked with eQTL data are green, and with both chromatin and eQTL are red.

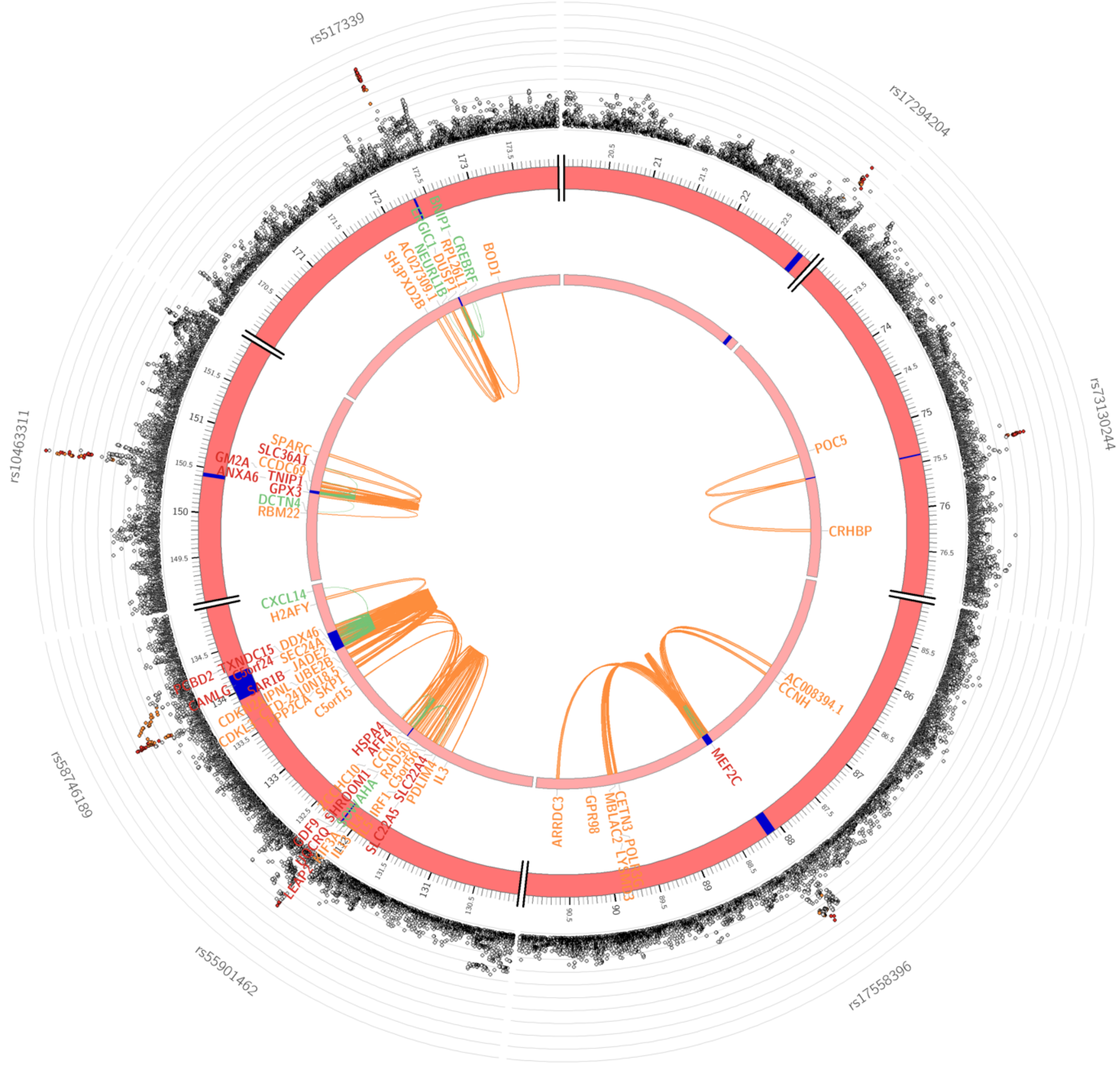


**Supplementary Figure 3**. **Circos plot of the Functional and Mapping annotation with cross-ancestry ALS GWAS summary statistics using FUMA on chromosome 5.** The outer dots indicate the -log$_{10}$ p-value with high LD SNPs with the lead SNP coloured red. Genes positionally linked with chromatin data are coloured orange, linked with eQTL data are green, and with both chromatin and eQTL are red.

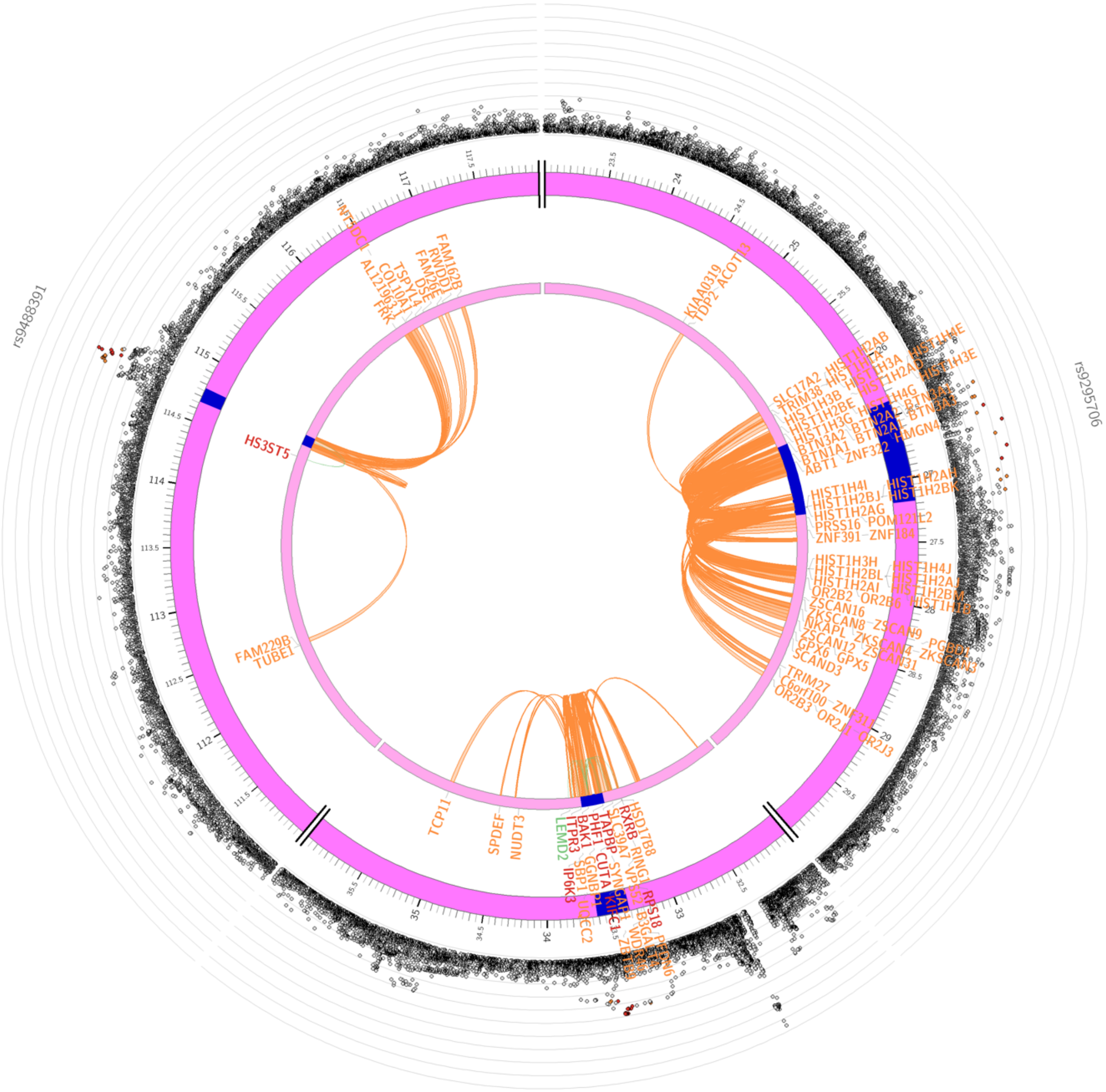


**Supplementary Figure 4**. **Circos plot of the Functional and Mapping annotation with cross-ancestry ALS GWAS summary statistics using FUMA on chromosome 6.** The outer dots indicate the -log$_{10}$ p-value with high LD SNPs with the lead SNP coloured red. Genes positionally linked with chromatin data are coloured orange, linked with eQTL data are green, and with both chromatin and eQTL are red.

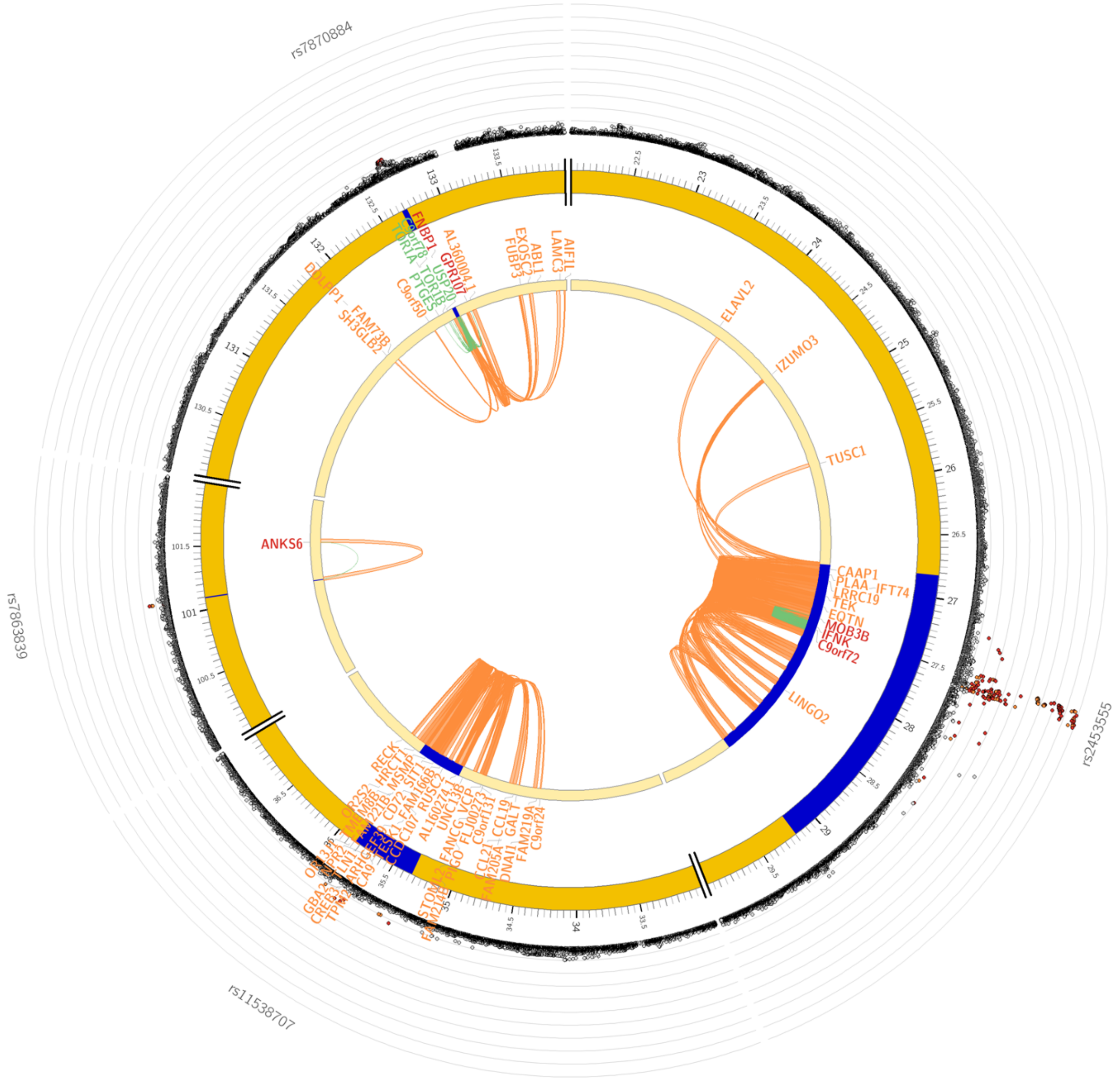


**Supplementary Figure 5**. **Circos plot of the Functional and Mapping annotation with cross-ancestry ALS GWAS summary statistics using FUMA on chromosome 9.** The outer dots indicate the -log10 p-value with high LD SNPs with the lead SNP coloured red. Genes positionally linked with chromatin data are coloured orange, linked with eQTL data are green, and with both chromatin and eQTL are red.

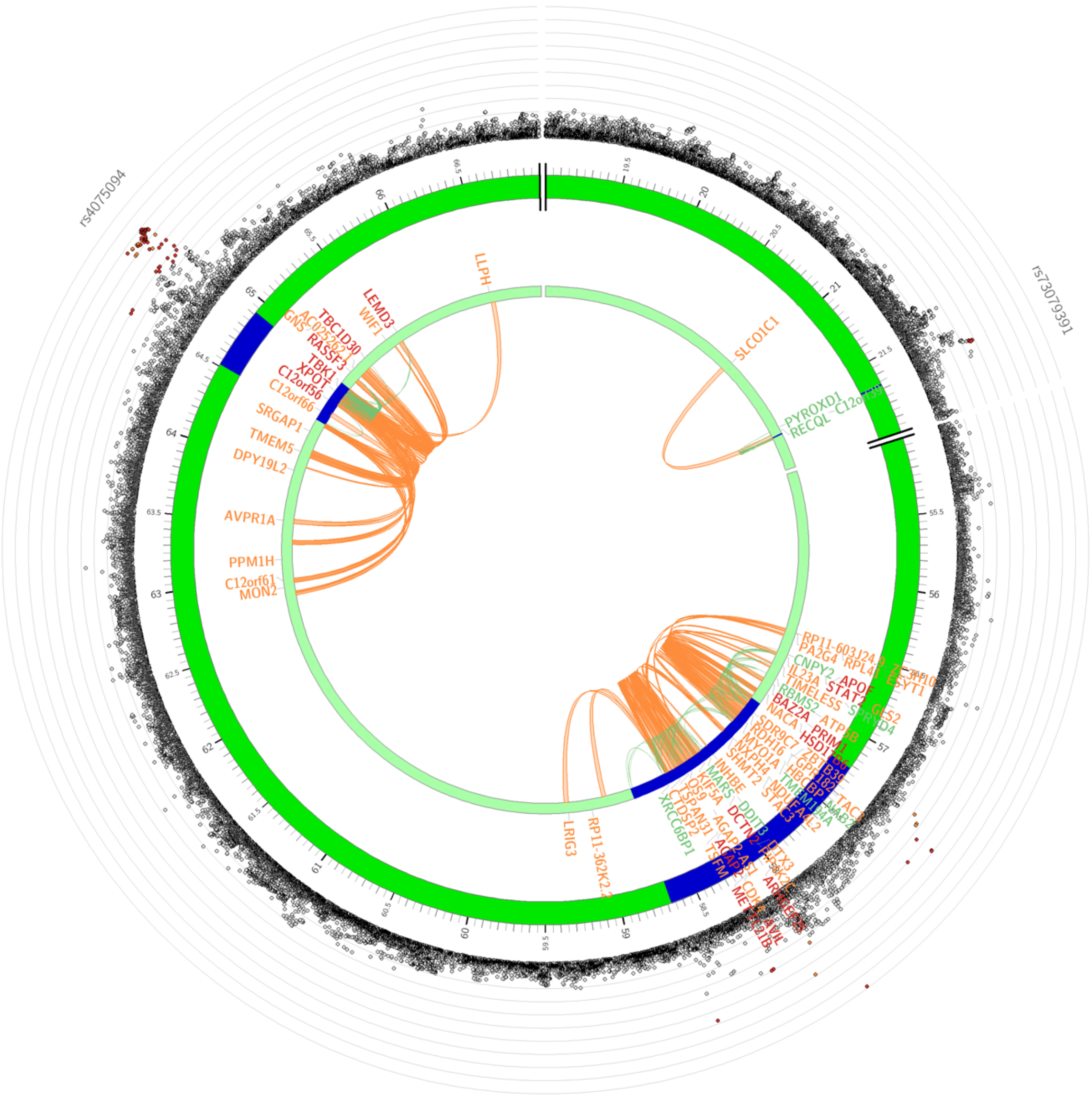

**Supplementary Figure 6**. **Circos plot of the Functional and Mapping annotation with cross-ancestry ALS GWAS summary statistics using FUMA on chromosome 12.** The outer dots indicate the -$\log_{10}$ p-value with high LD SNPs with the lead SNP coloured red. Genes positionally linked with chromatin data are coloured orange, linked with eQTL data are green, and with both chromatin and eQTL are red.

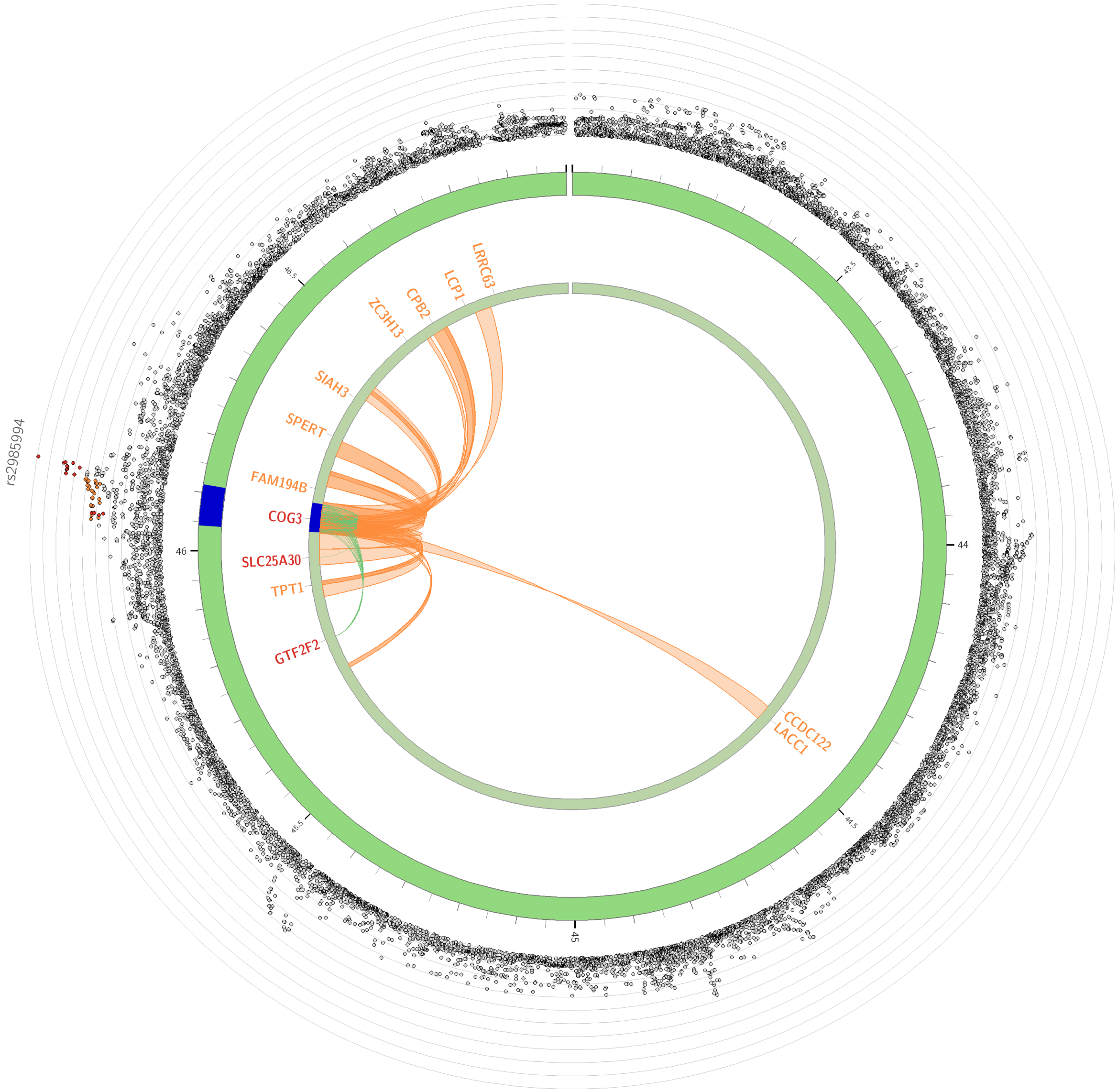


**Supplementary Figure 7**. **Circos plot of the Functional and Mapping annotation with cross-ancestry ALS GWAS summary statistics using FUMA on chromosome 13.** The outer dots indicate the $-\log_{10}$ p-value with high LD SNPs with the lead SNP coloured red. Genes positionally linked with chromatin data are coloured orange, linked with eQTL data are green, and with both chromatin and eQTL are red.

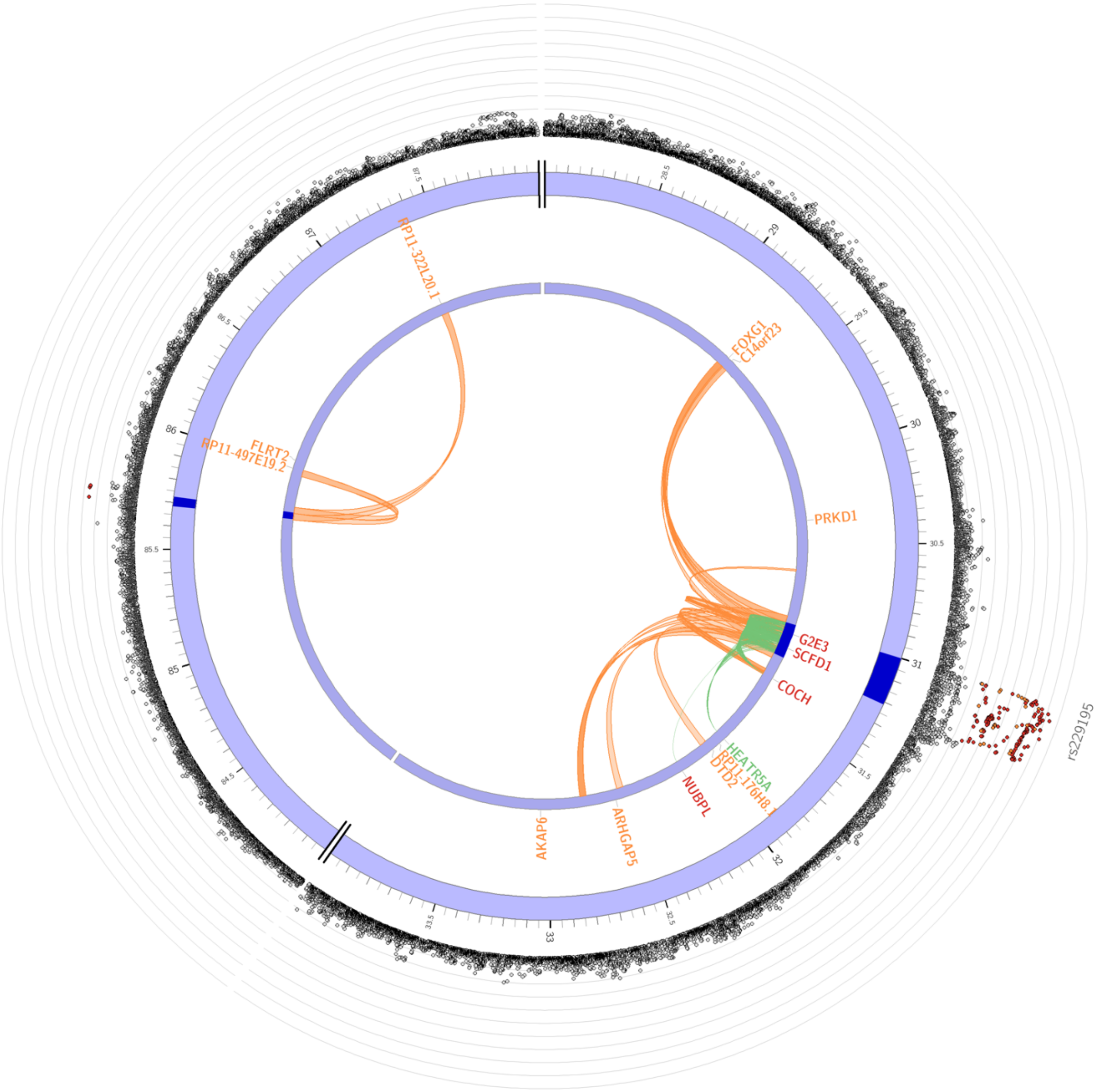


**Supplementary Figure 8**. **Circos plot of the Functional and Mapping annotation with cross-ancestry ALS GWAS summary statistics using FUMA on chromosome 14.** The outer dots indicate the -$\log_{10}$ p-value with high LD SNPs with the lead SNP coloured red. Genes positionally linked with chromatin data are coloured orange, linked with eQTL data are green, and with both chromatin and eQTL are red.

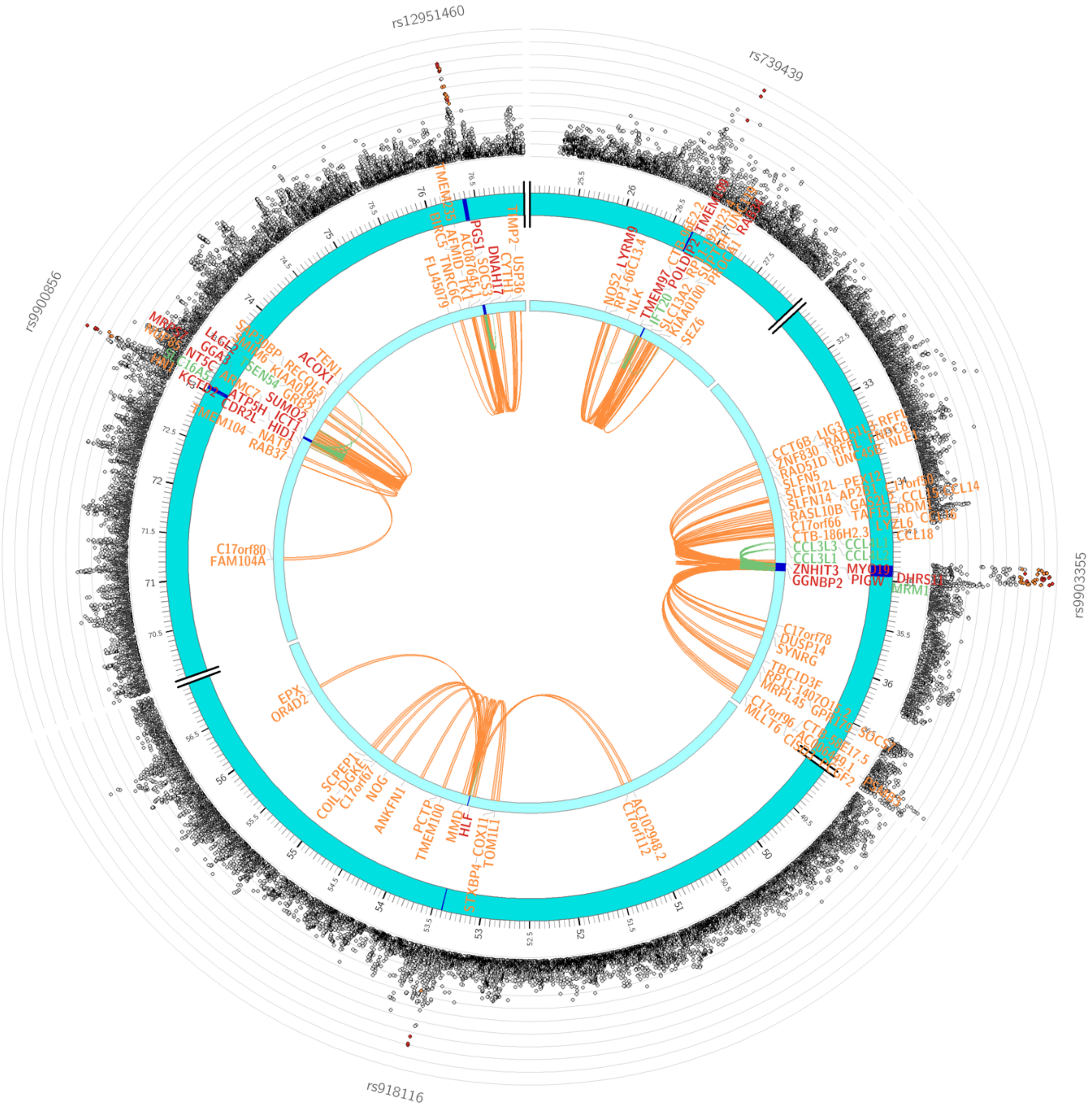


**Supplementary Figure 9**. **Circos plot of the Functional and Mapping annotation with cross-ancestry ALS GWAS summary statistics using FUMA on chromosome 17.** The outer dots indicate the -log$_{10}$ p-value with high LD SNPs with the lead SNP coloured red. Genes positionally linked with chromatin data are coloured orange, linked with eQTL data are green, and with both chromatin and eQTL are red.

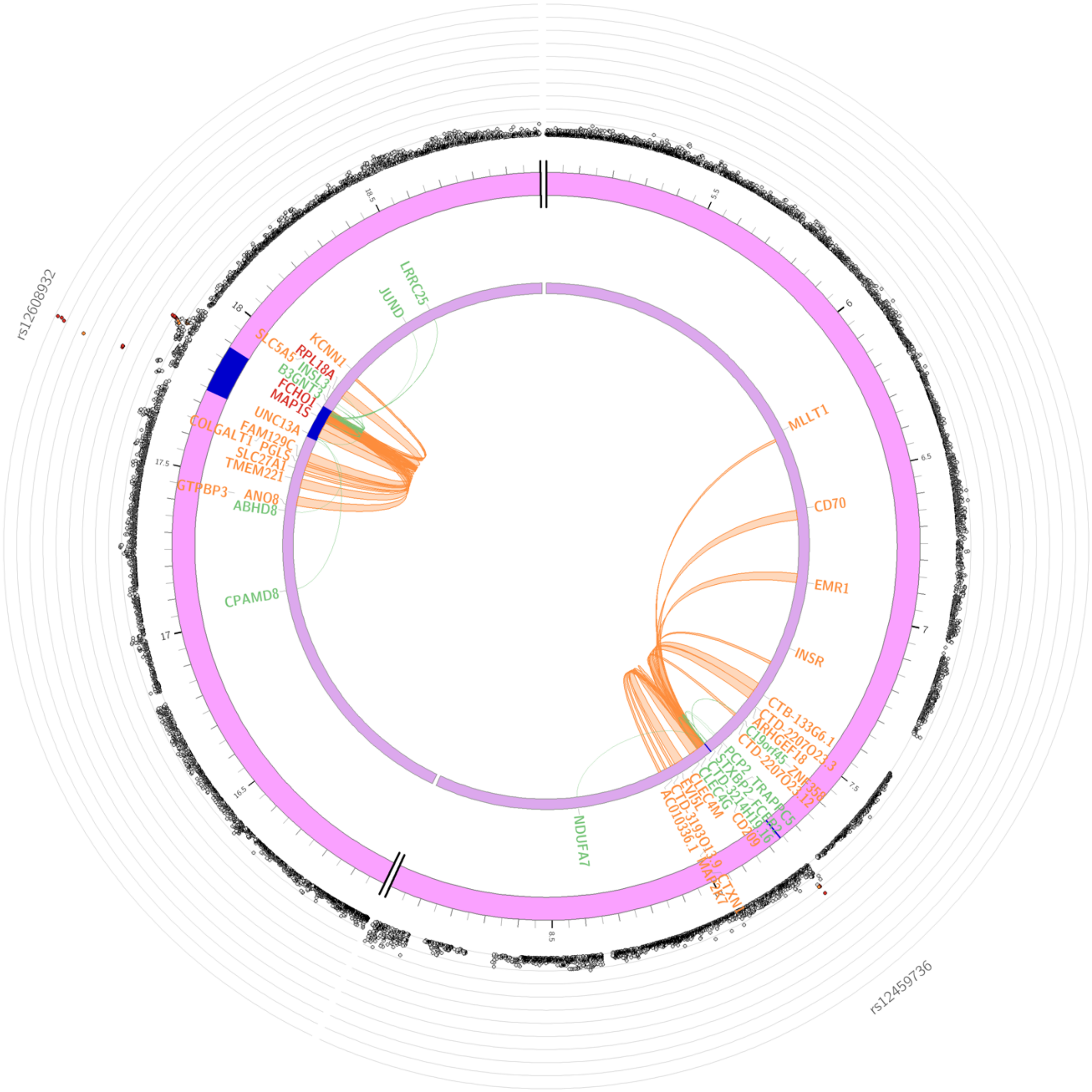

**Supplementary Figure 10**. **Circos plot of the Functional and Mapping annotation with cross-ancestry ALS GWAS summary statistics using FUMA on chromosome 19.** The outer dots indicate the $-\log_{10}$ p-value with high LD SNPs with the lead SNP coloured red. Genes positionally linked with chromatin data are coloured orange, linked with eQTL data are green, and with both chromatin and eQTL are red.

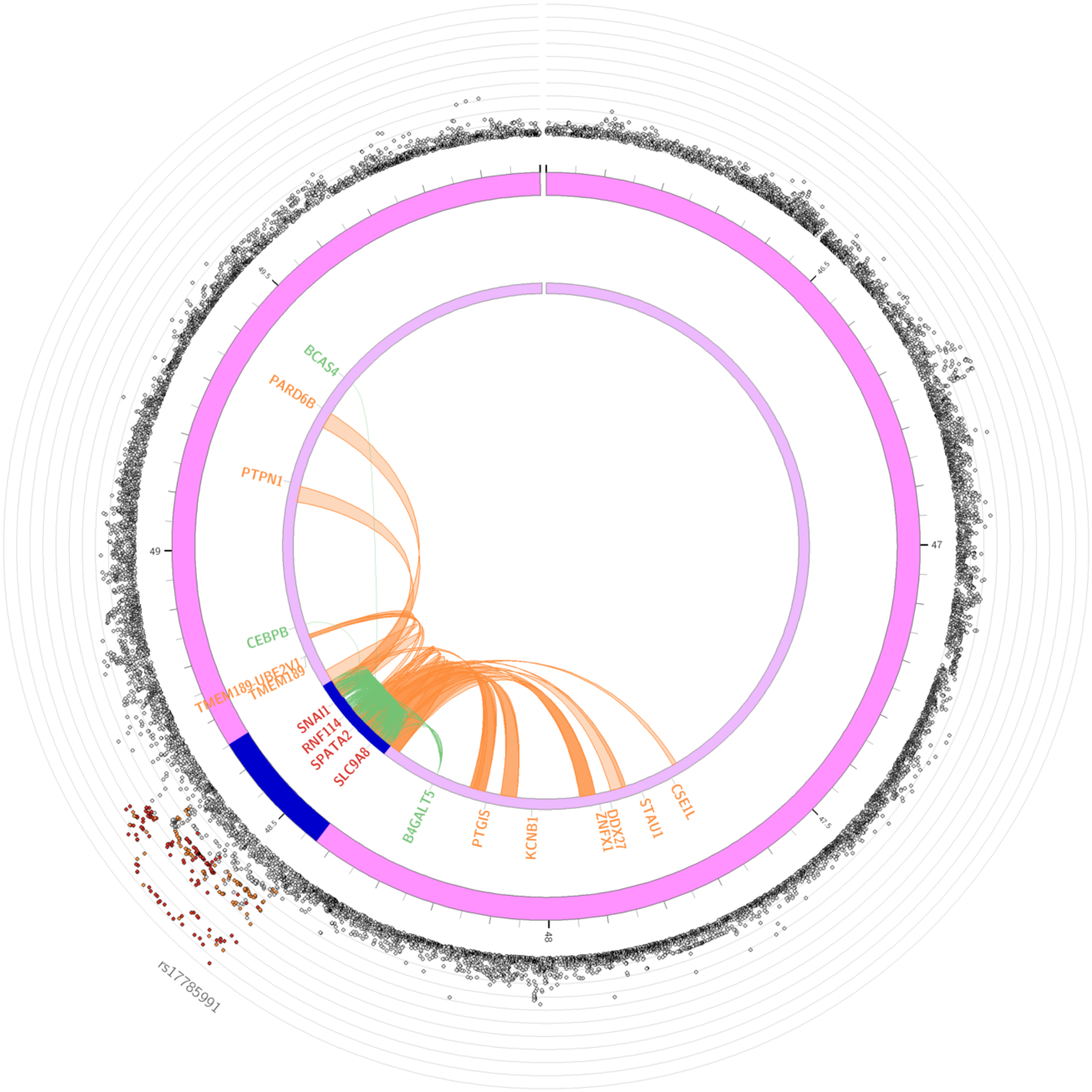


**Supplementary Figure 11**. **Circos plot of the Functional and Mapping annotation with cross-ancestry ALS GWAS summary statistics using FUMA on chromosome 20.** The outer dots indicate the $-\log_{10}$ p-value with high LD SNPs with the lead SNP coloured red. Genes positionally linked with chromatin data are coloured orange, linked with eQTL data are green, and with both chromatin and eQTL are red.

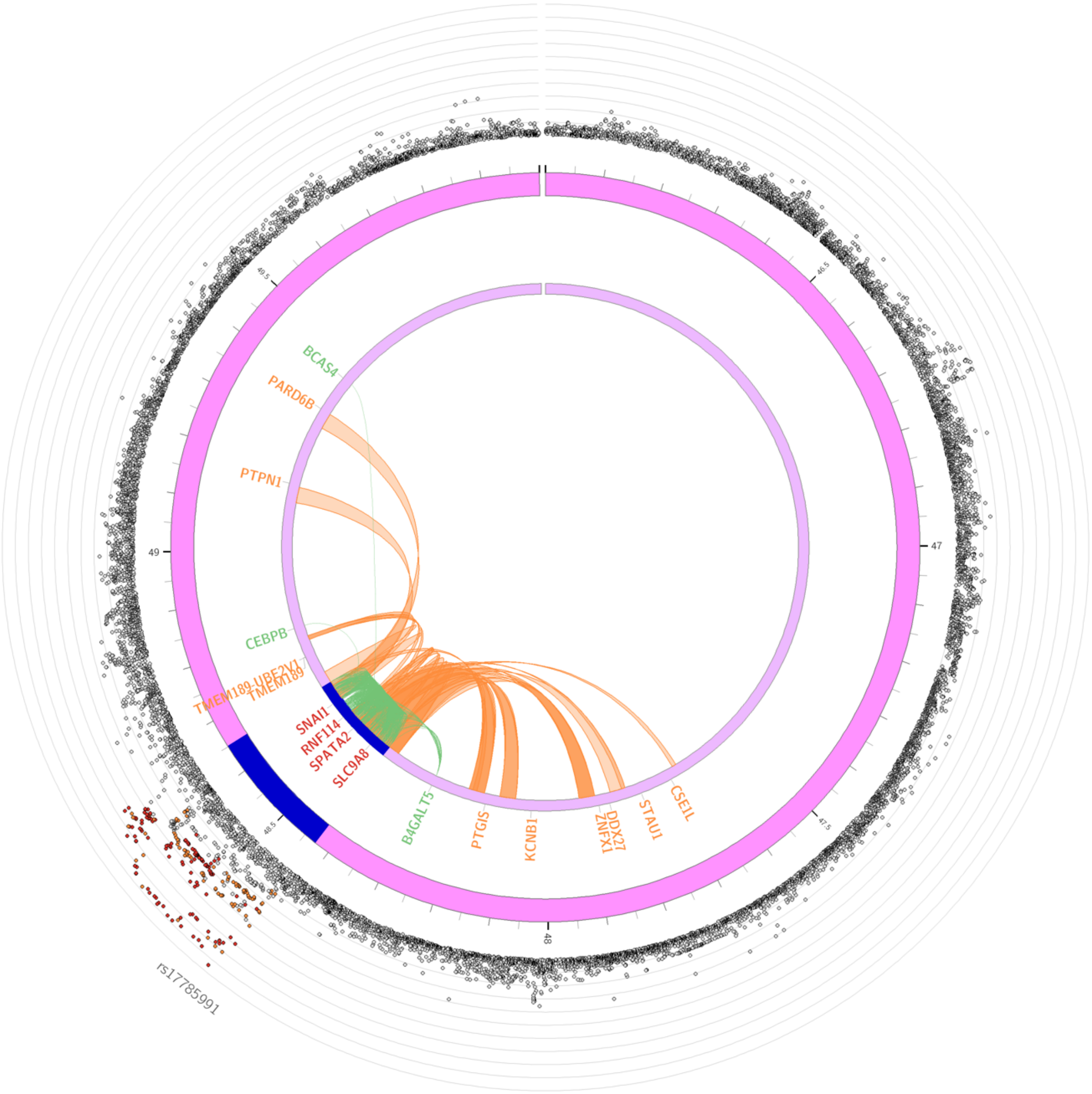


**Supplementary Figure 12**. **Circos plot of the Functional and Mapping annotation with cross-ancestry ALS GWAS summary statistics using FUMA on chromosome 20.** The outer dots indicate the $-\log_{10}$ p-value with high LD SNPs with the lead SNP coloured red. Genes positionally linked with chromatin data are coloured orange, linked with eQTL data are green, and with both chromatin and eQTL are red.

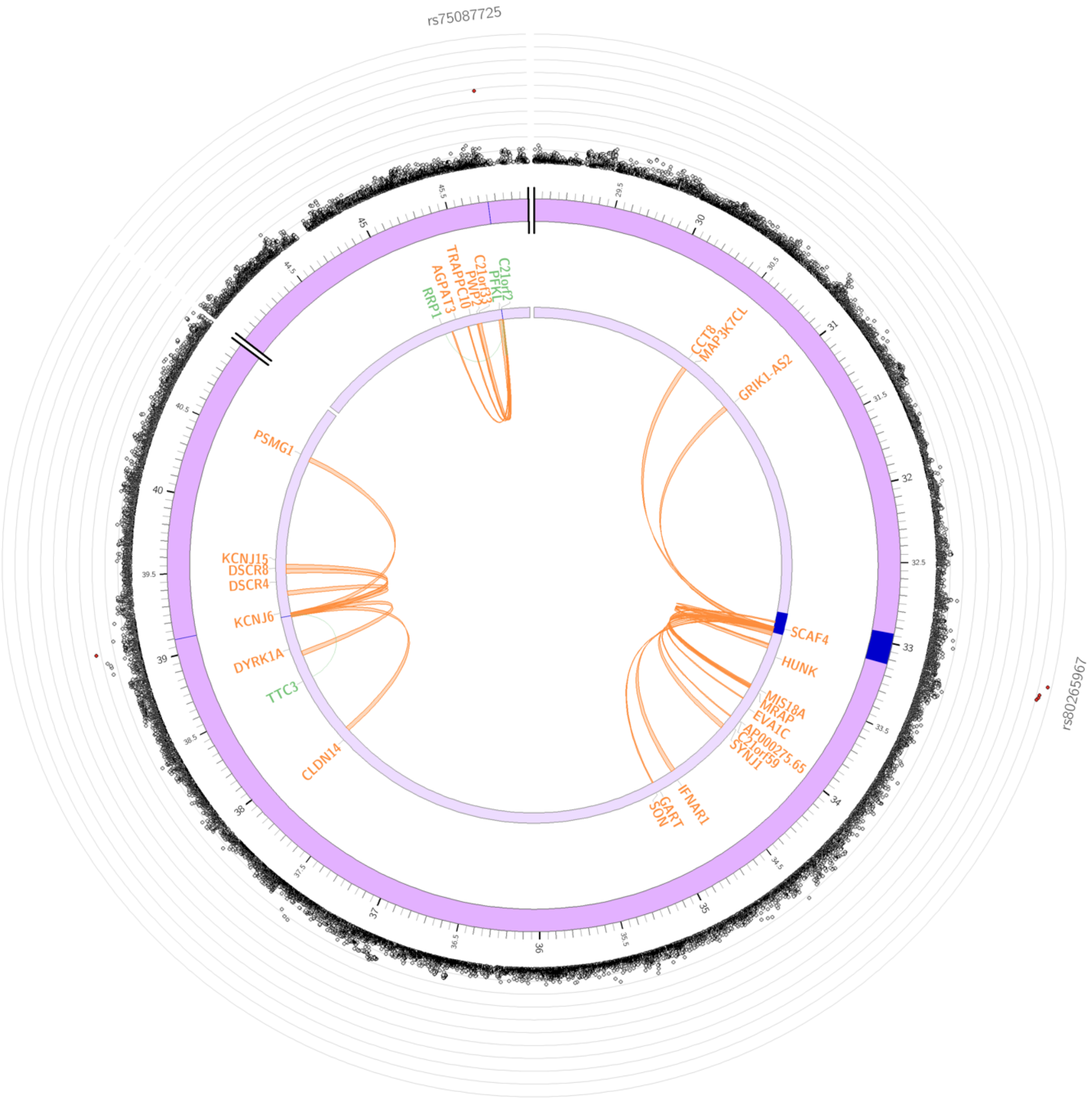


**Supplementary Figure 13**. **Circos plot of the Functional and Mapping annotation with cross-ancestry ALS GWAS summary statistics using FUMA on chromosome 21.** The outer dots indicate the $-\log_{10}$ p-value with high LD SNPs with the lead SNP coloured red. Genes positionally linked with chromatin data are coloured orange, linked with eQTL data are green, and with both chromatin and eQTL are red.

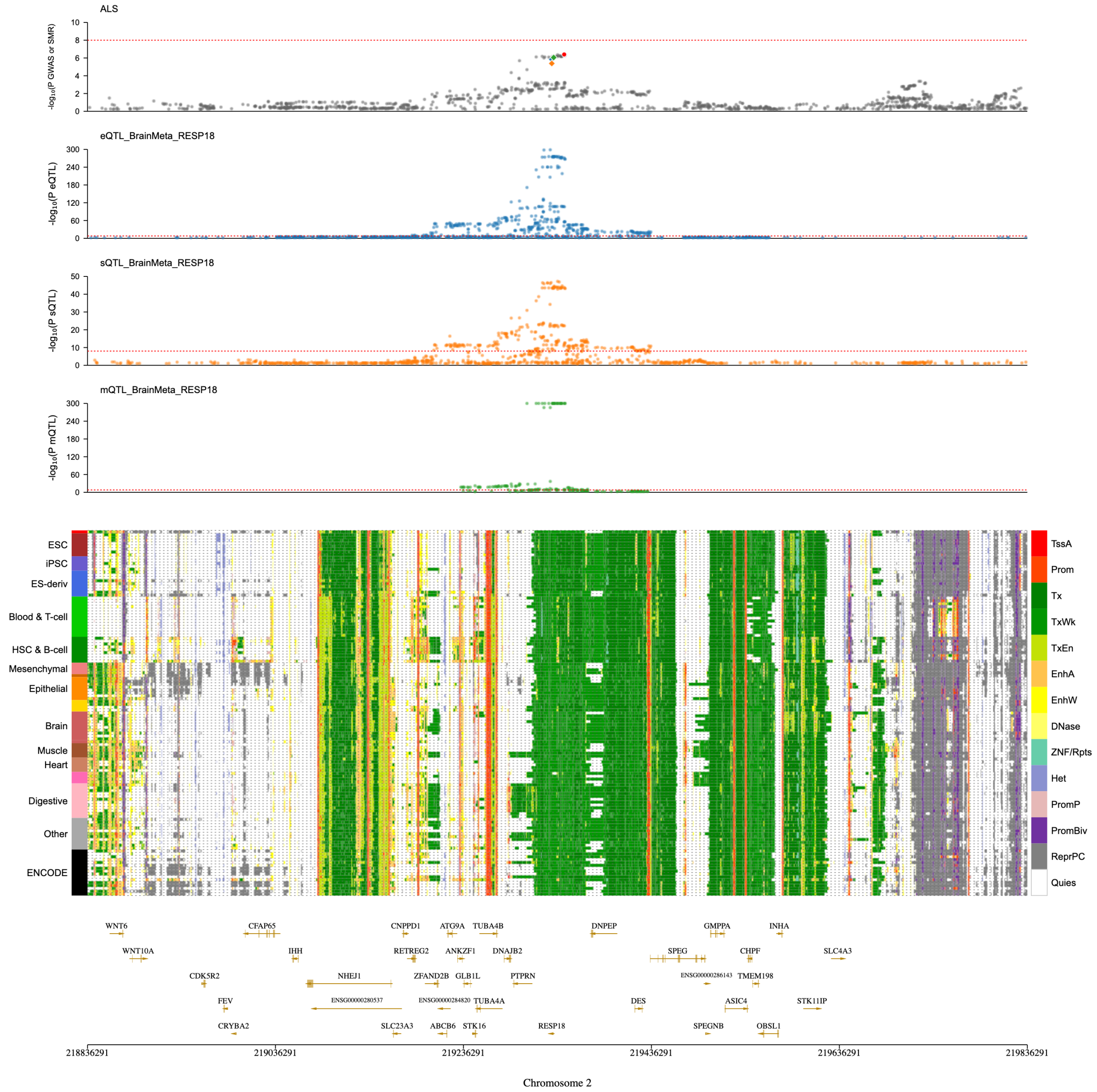


**Supplementary Figure 14**. **Significant SMR results on chromosome 2.** A) ALS GWAS/SMR plot. Followed by eQTL (blue), sQTL (orange), mQTL (green). The red dashed line indicates *p* value threshold of $< 5 \times 10^{-8}$. B) Epigenomic annotation of cells/tissues with corresponding gene map.

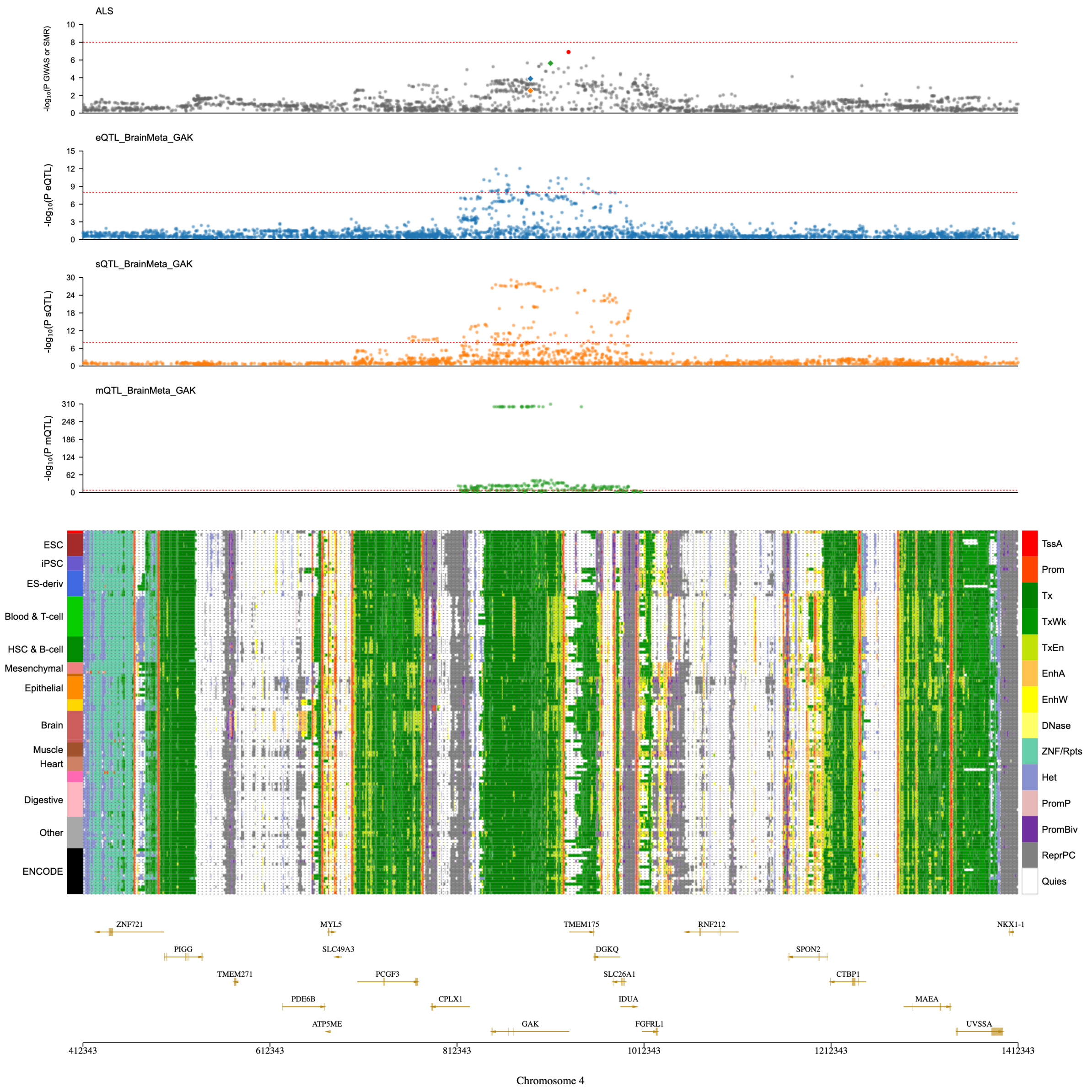


**Supplementary Figure 15**. **Significant SMR results on chromosome 4.** A) ALS GWAS/SMR plot, followed by eQTL (blue), sQTL (orange), mQTL (green). The red dashed line indicates *p* value threshold of $< 5 \times 10^{-8}$. B) Epigenomic annotation of cells/tissues with corresponding gene map.

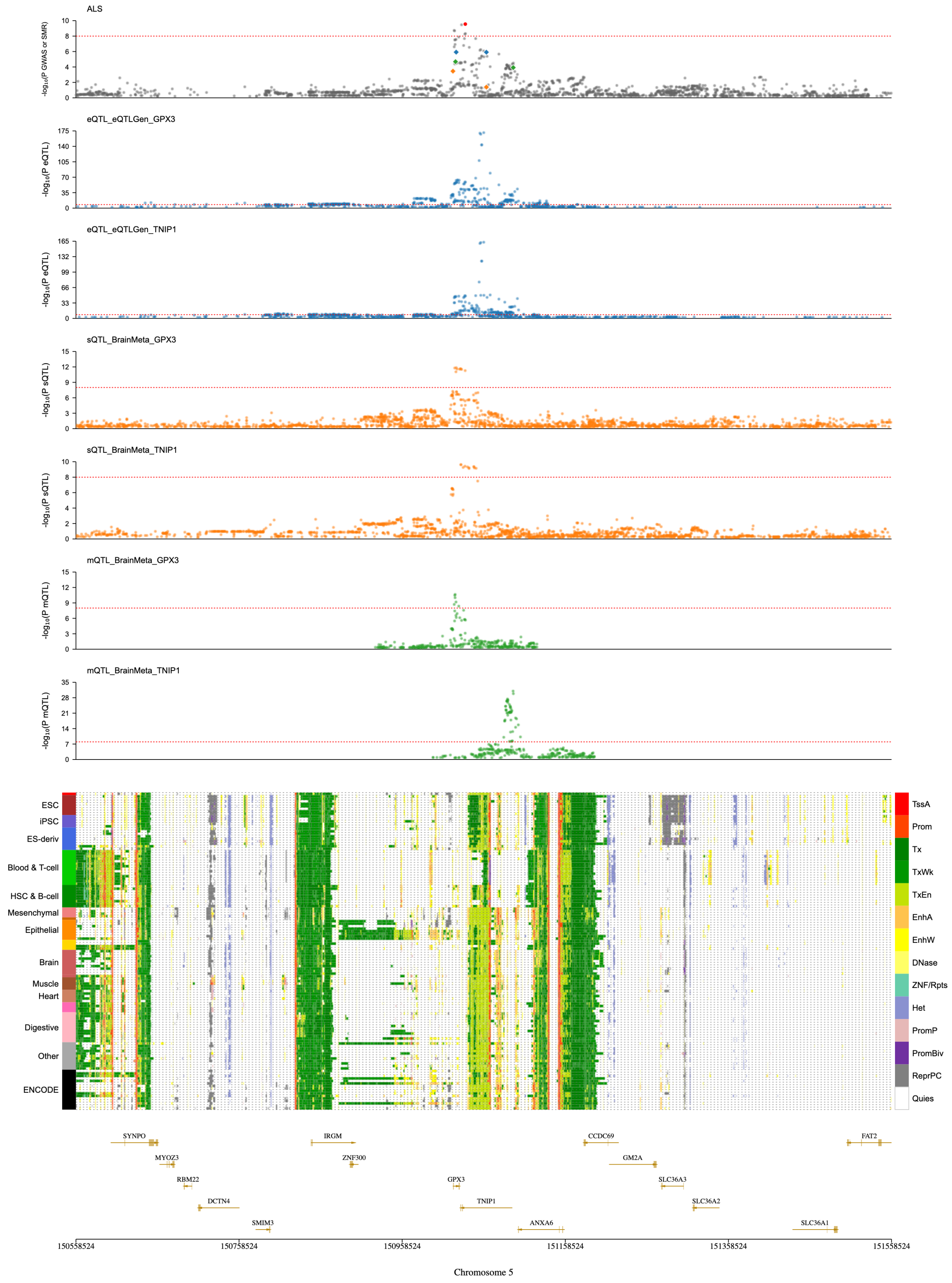


**Supplementary Figure 16**. **Significant SMR results on chromosome 5.** A) ALS GWAS/SMR plot, followed by eQTL (blue), sQTL (orange), mQTL (green). The red dashed line indicates *p* value threshold of $< 5 \times 10^{-8}$. B) Epigenomic annotation of cells/tissues with corresponding gene map.

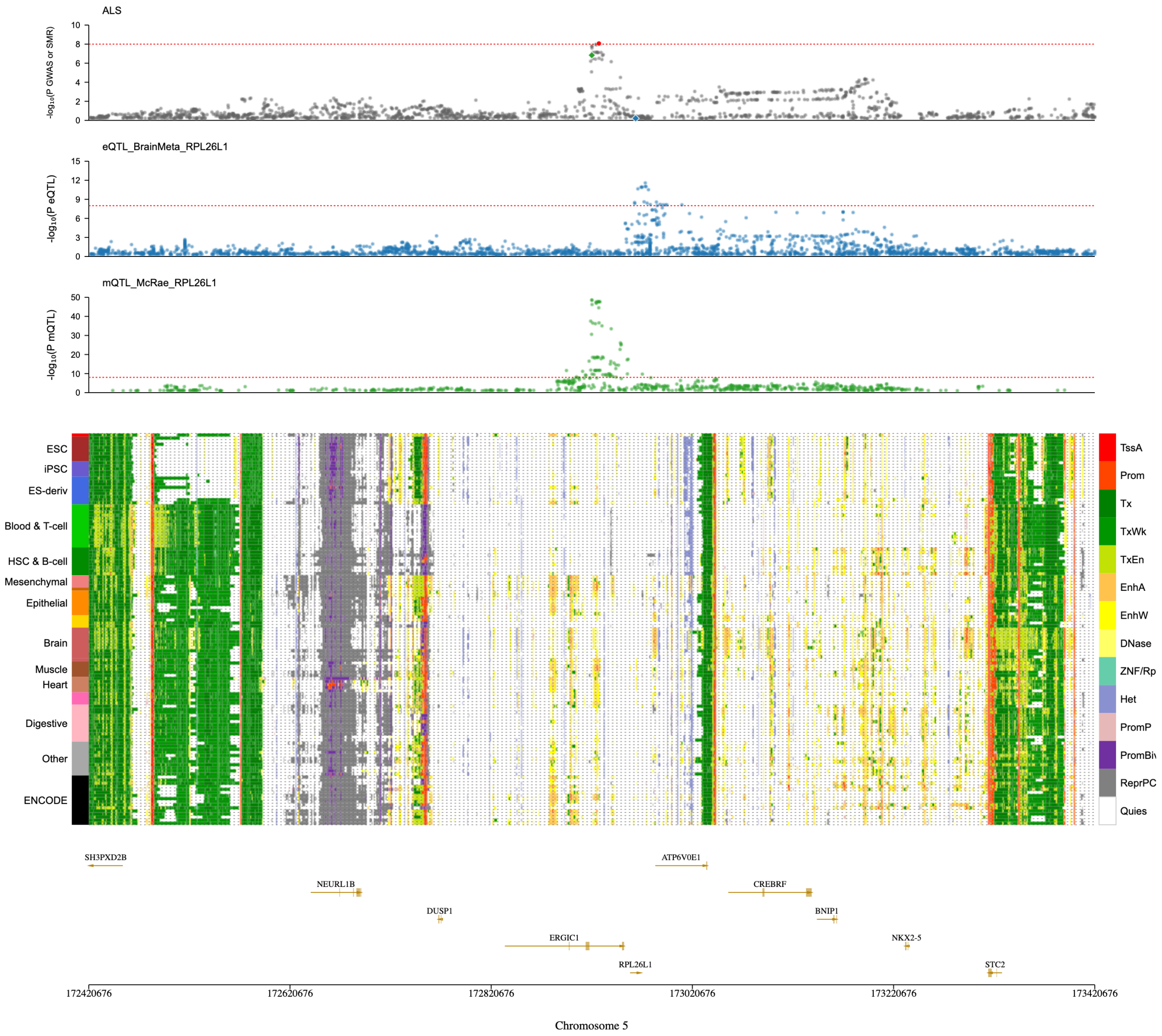


**Supplementary Figure 17**. **Significant SMR results on chromosome 5.** A) ALS GWAS/SMR plot, followed by eQTL (blue), sQTL (orange), mQTL (green). The red dashed line indicates *p* value threshold of $< 5 \times 10^{-8}$. B) Epigenomic annotation of cells/tissues with corresponding gene map.

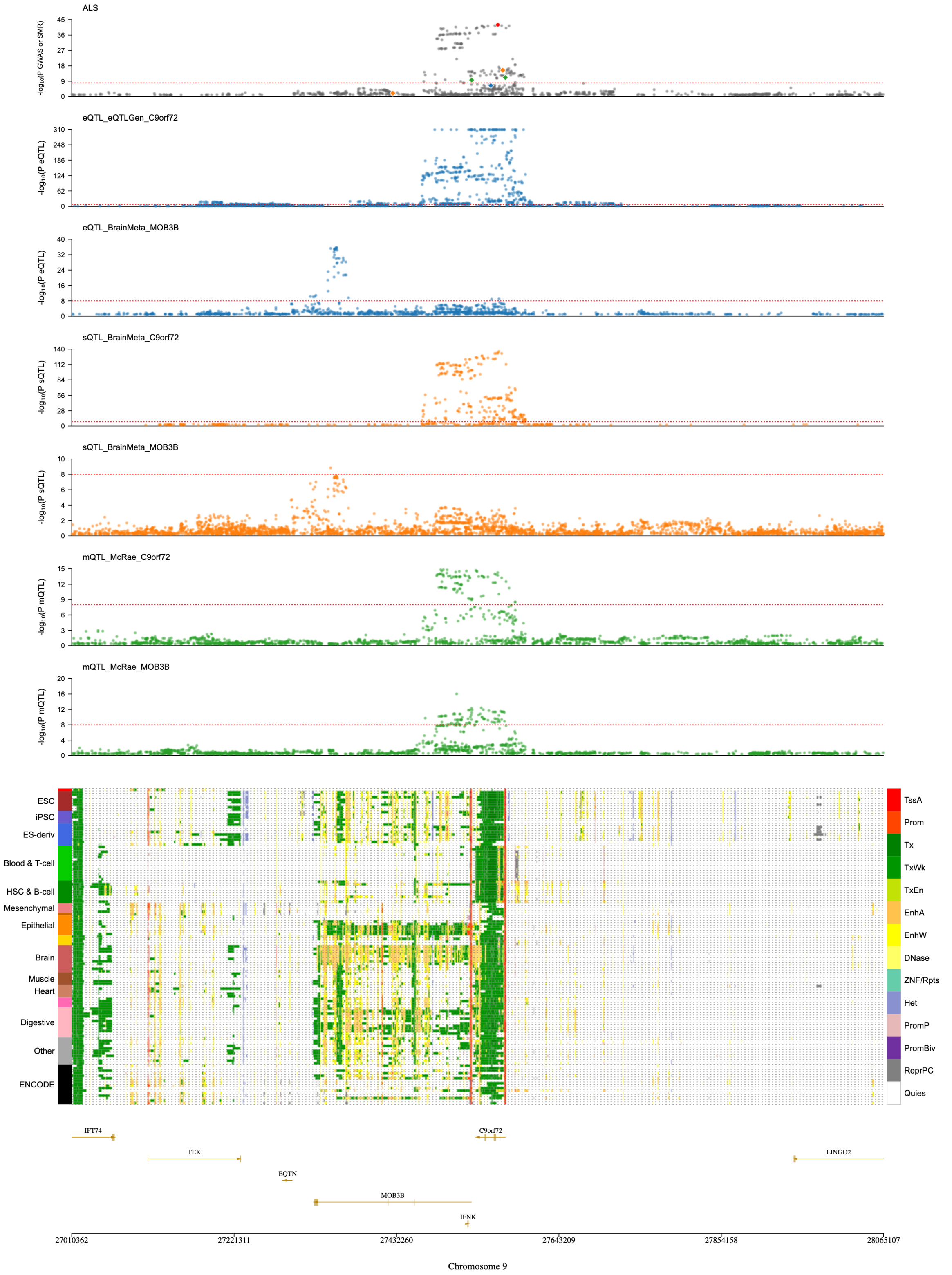


**Supplementary Figure 18**. **Significant SMR results on chromosome 9.** A) ALS GWAS/SMR plot, followed by eQTL (blue), sQTL (orange), mQTL (green). The red dashed line indicates *p* value threshold of $< 5 \times 10^{-8}$. B) Epigenomic annotation of cells/tissues with corresponding gene map.

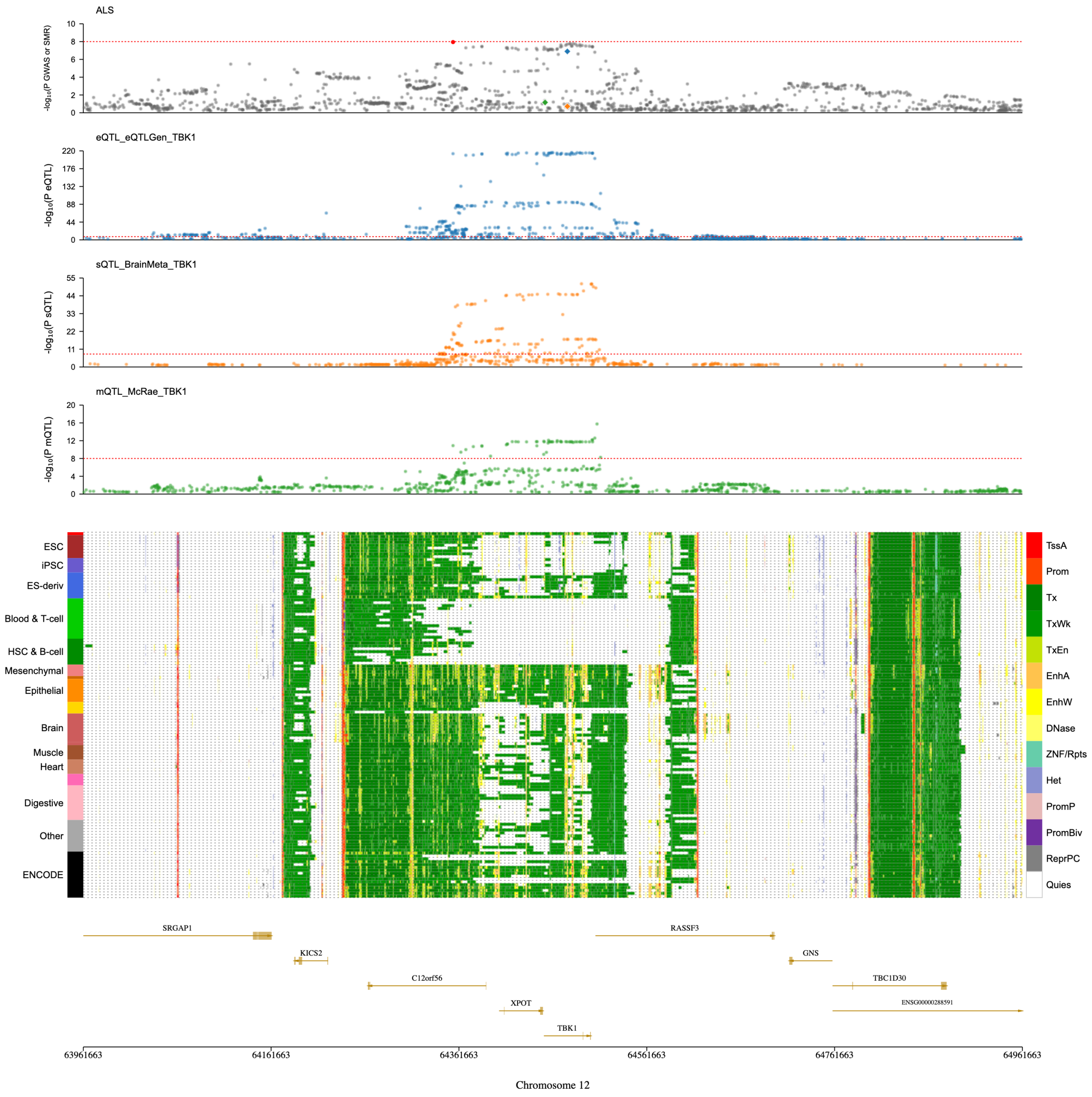


**Supplementary Figure 19**. **Significant SMR results on chromosome 12.** A) ALS GWAS/SMR plot, followed by eQTL (blue), sQTL (orange), mQTL (green). The red dashed line indicates $p$ value threshold of $< 5 \times 10^{-8}$. B) Epigenomic annotation of cells/tissues with corresponding gene map.

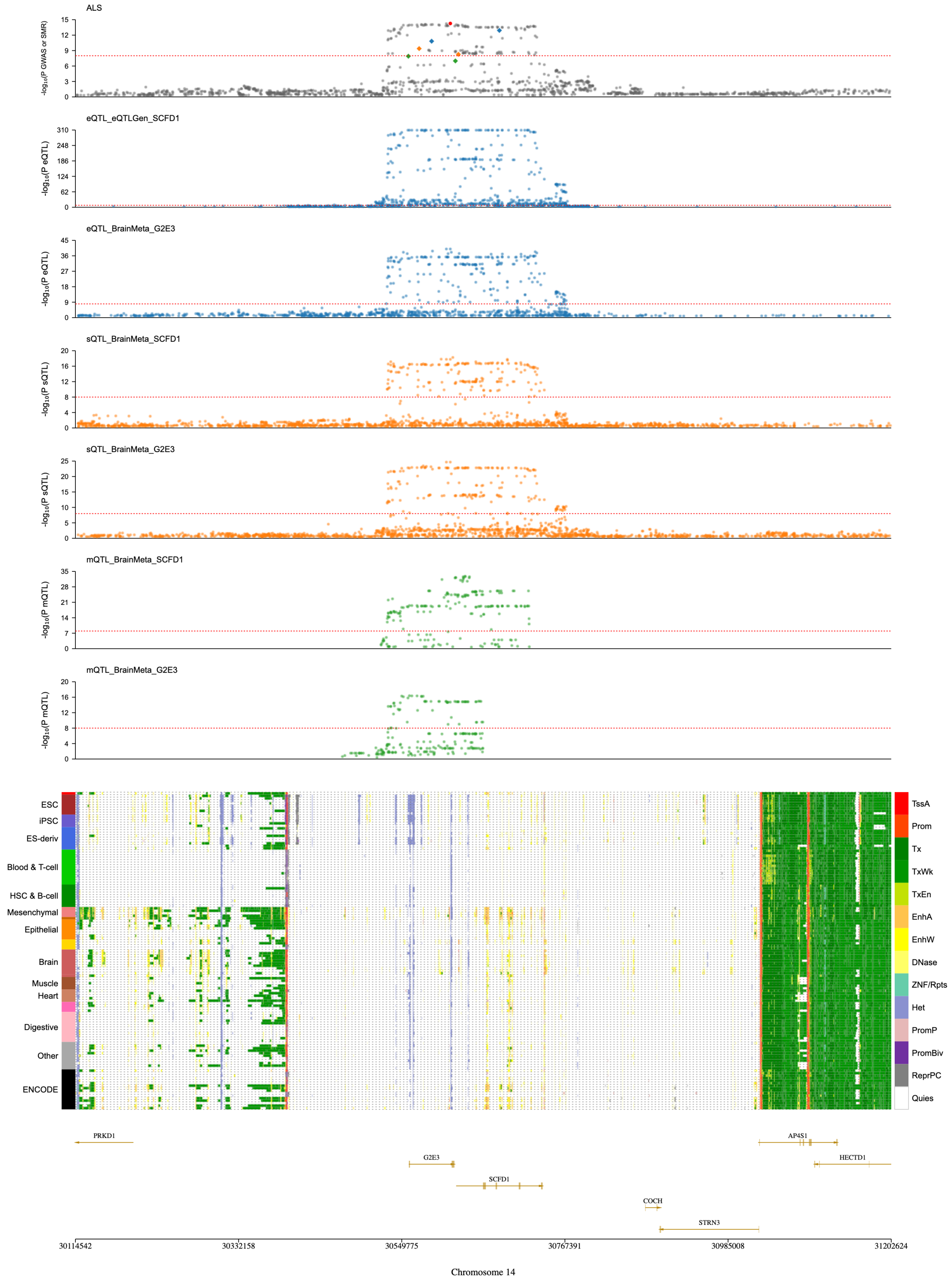


**Supplementary Figure 20**. **Significant SMR results on chromosome 14.** A) ALS GWAS/SMR plot, followed by eQTL (blue), sQTL (orange), mQTL (green). The red dashed line indicates *p* value threshold of $< 5 \times 10^{-8}$. B) Epigenomic annotation of cells/tissues with corresponding gene map.

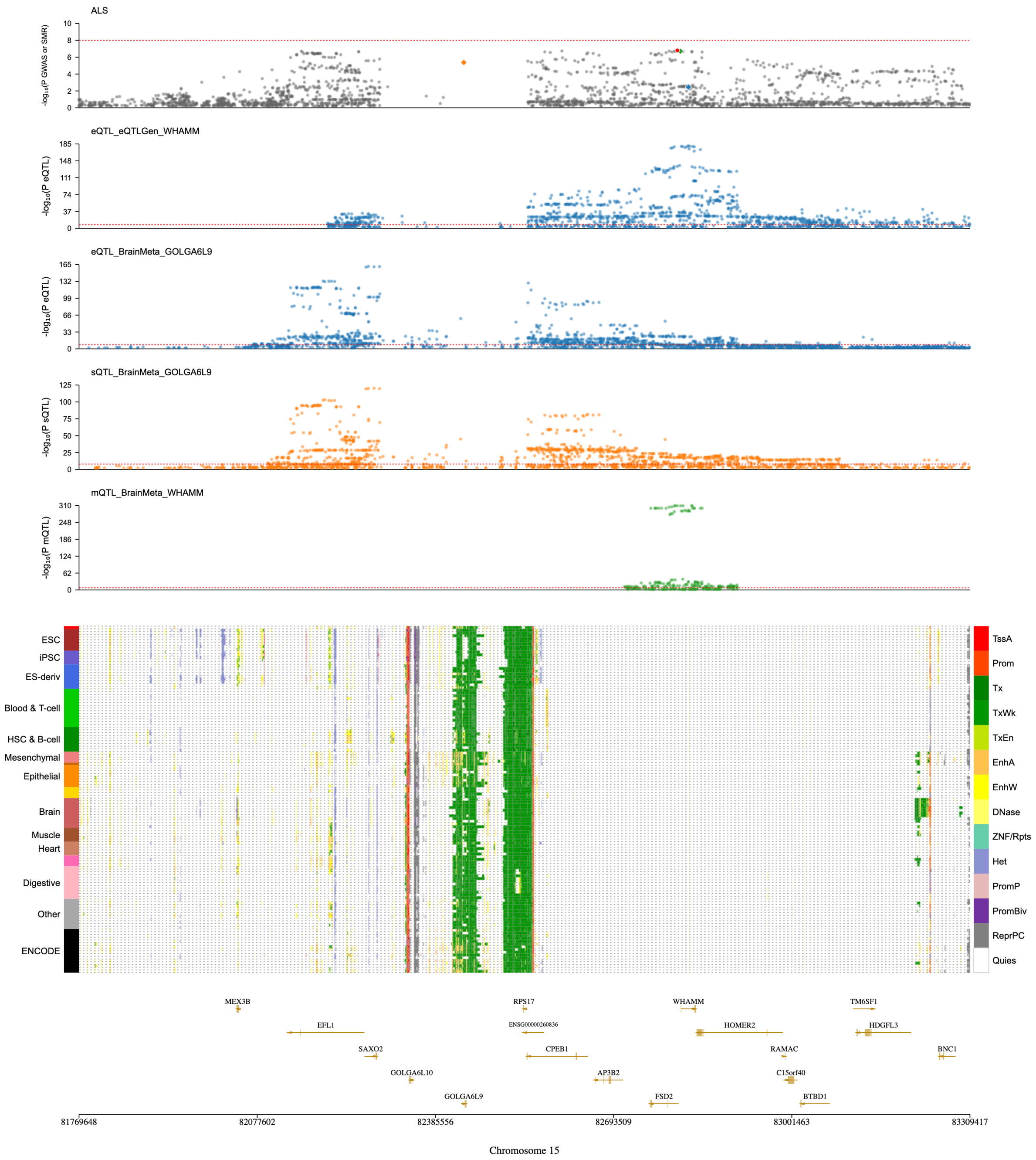


**Supplementary Figure 21**. **Significant SMR results on chromosome 15.** A) ALS GWAS/SMR plot, followed by eQTL (blue), sQTL (orange), mQTL (green). The red dashed line indicates *p* value threshold of $< 5 \times 10^{-8}$. B) Epigenomic annotation of cells/tissues with corresponding gene map.

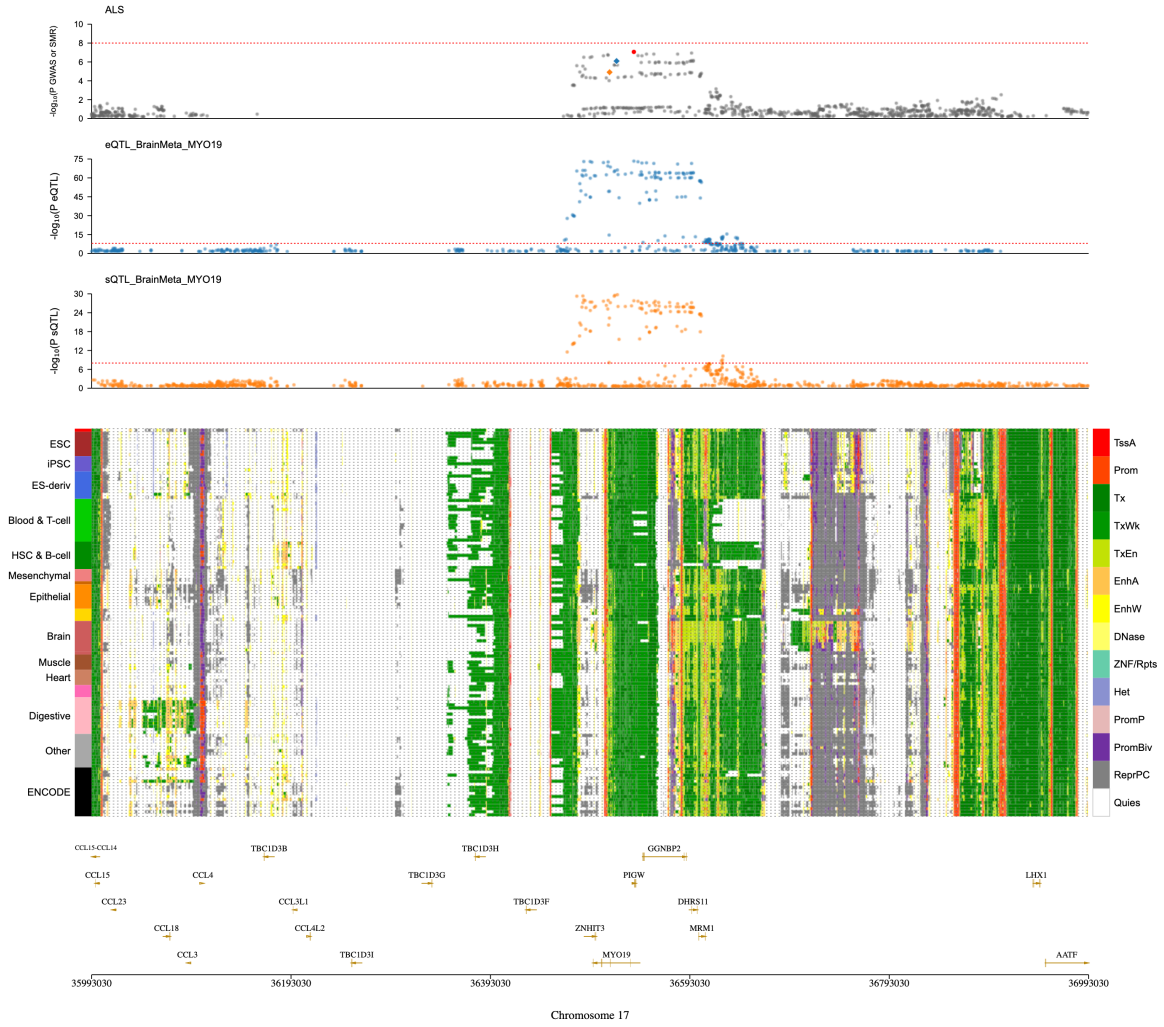


**Supplementary Figure 22**. **Significant SMR results on chromosome 17.** A) ALS GWAS/SMR plot, followed by eQTL (blue) and sQTL (orange). The red dashed line indicates $p$ value threshold of $< 5 \times 10^{-8}$. B) Epigenomic annotation of cells/tissues with corresponding gene map.

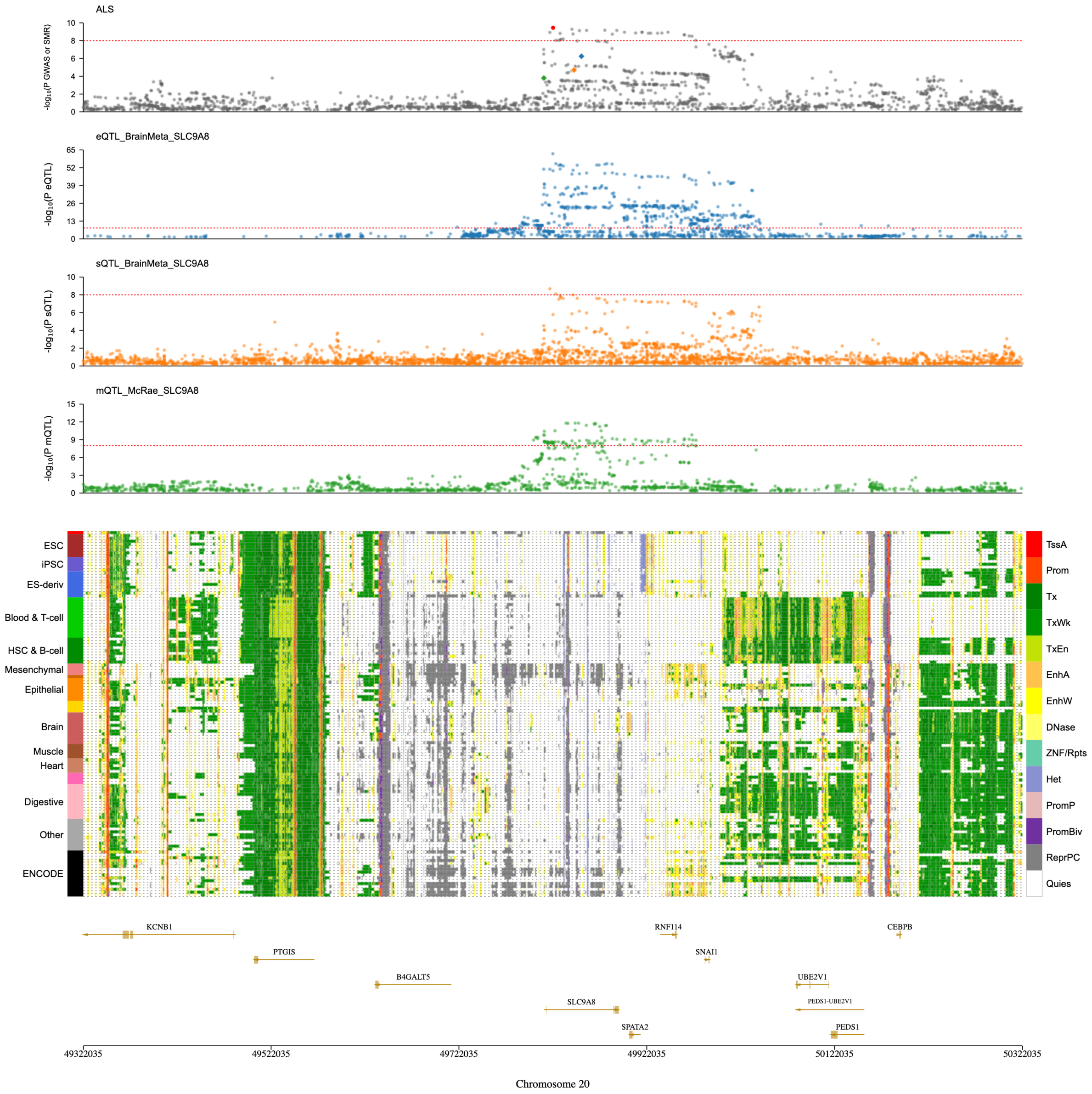


**Supplementary Figure 23**. **Significant SMR results on chromosome 20.** A) ALS GWAS/SMR plot, followed by eQTL (blue), sQTL (orange), mQTL (green). The red dashed line indicates *p* value threshold of $< 5 \times 10^{-8}$. B) Epigenomic annotation of cells/tissues with corresponding gene map.

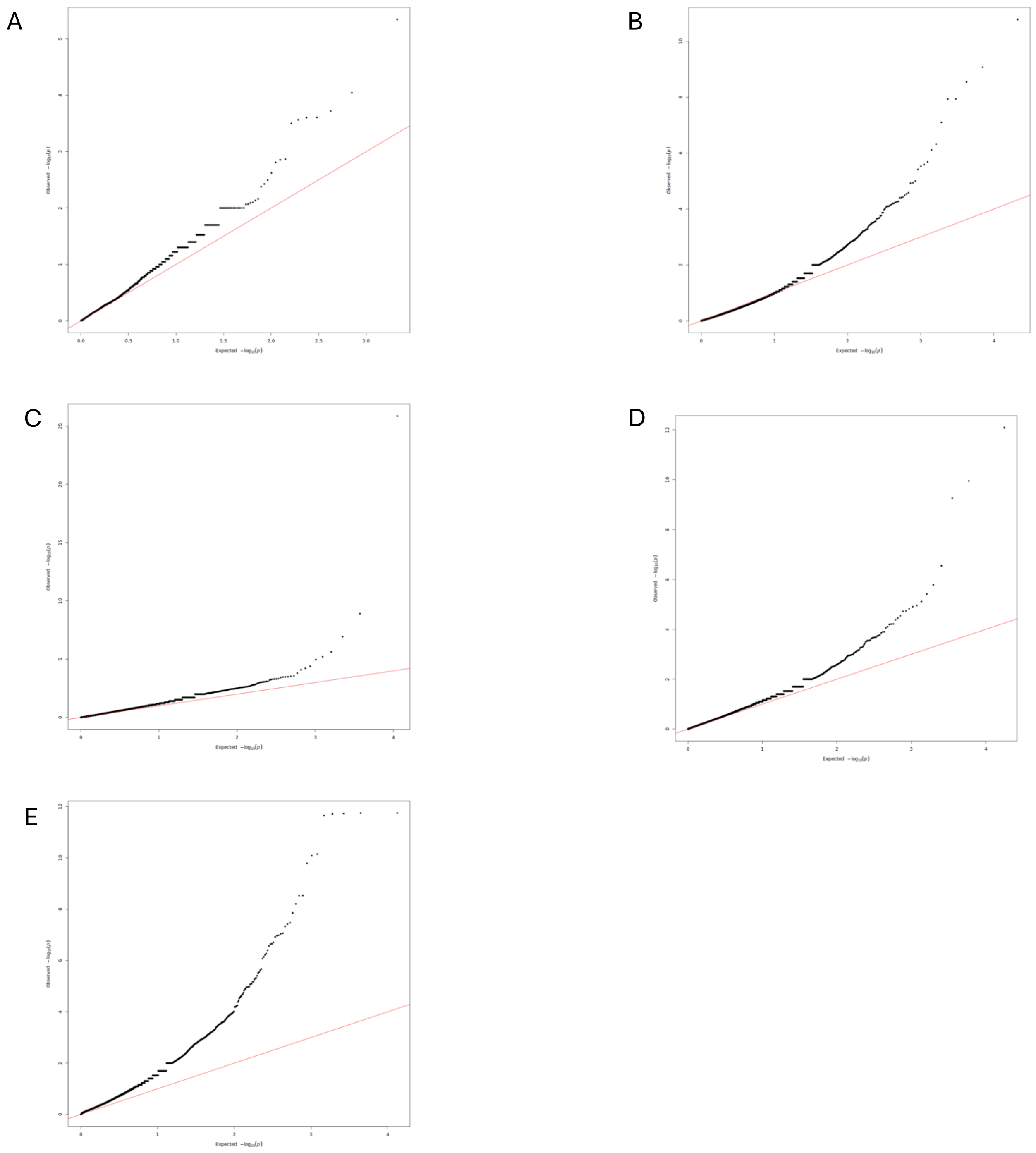


**Supplementary Figure 24**. **TWAS QQ plots.** A) FUSION B) GIFT C) S-PrediXcan D) TF-TWAS E) UTMOST. Most methods have minimal inflation, except for UTMOST.

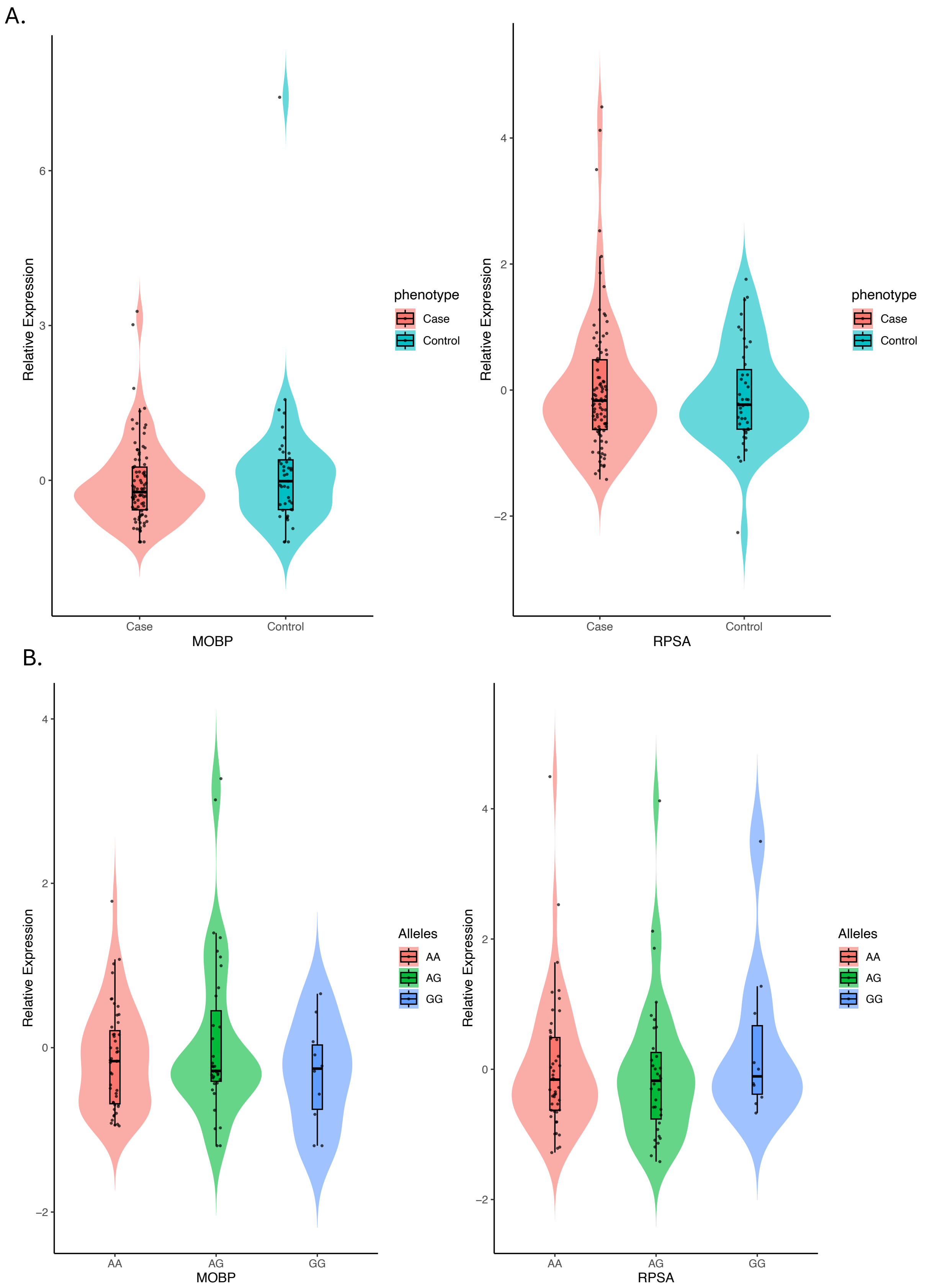


**Supplementary Figure 25. Blood RNA expression in ALS cases and controls. A)** *RPSA* or *MOBP* expression between ALS cases and controls (*p*=0.64 and 0.39, respectively, n=177). B) For samples that had matched genotype data (n=135), no associations were detected with lead risk SNP (rs631312, A=0.67, G=0.33) and *RPSA* or *MOBP* gene expression using a linear expression model.

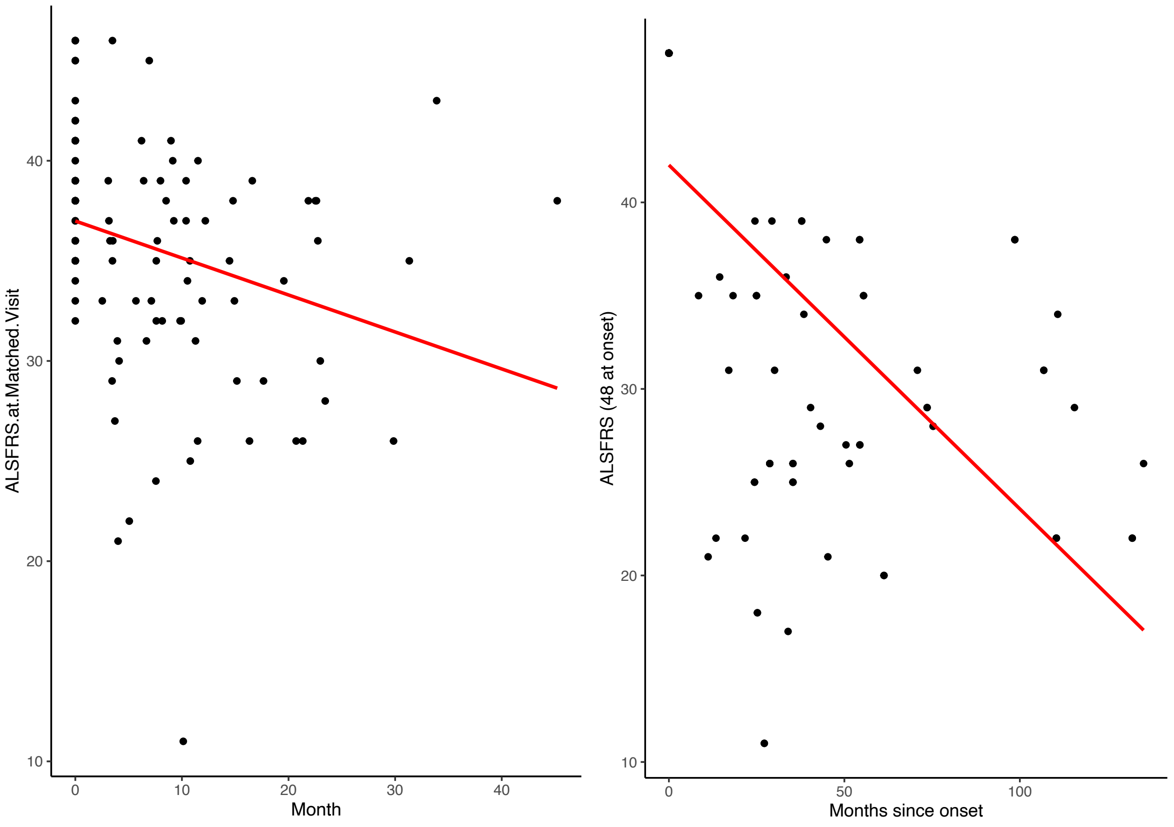


**Supplementary Figure 26.** Longitudinal Australian ALS case cohort (n=41 individuals, n=103 observations, 2-4 visits). The change in ALSFRS-R in months since first visit and months since diagnosis we identified a linear decrease of 0.18 points (-0.31 to -0.06 95% CI, $p$=0.004) and 0.18 (-0.23 to -0.14 95% CI, $p$=3.51e-12) each month respectively.

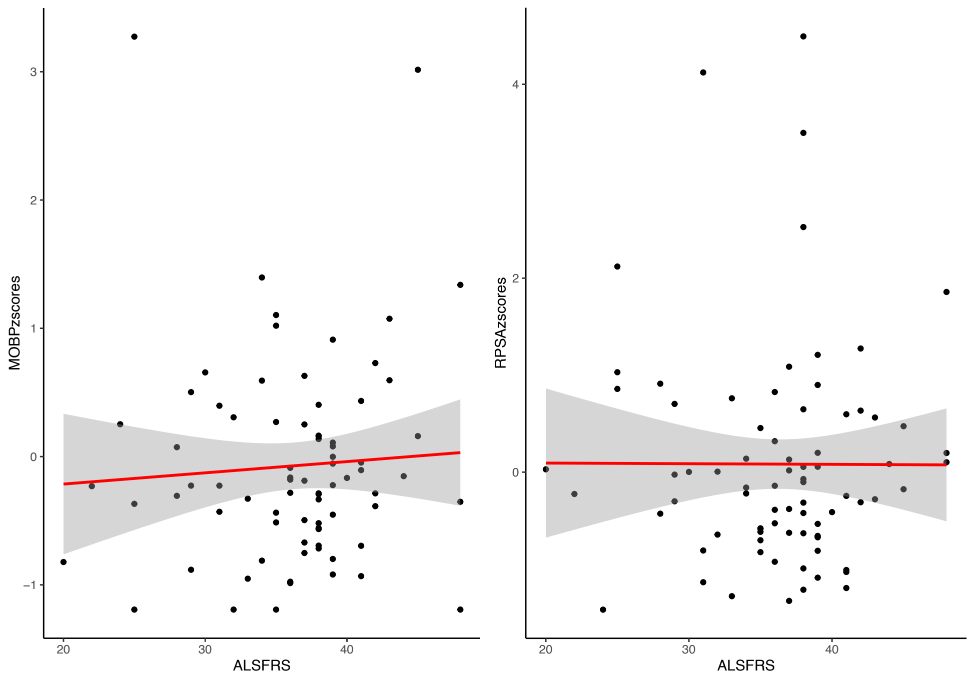


**Supplementary Figure 27**. Using a linear mixed-model analysis, fitting individual as a random effect, there was no association with *MOBP* or *RPSA* expression and ALS functional rating score (ALSFRS-R (scale from 48 to 0, where 48 is normal physical function) was identified.

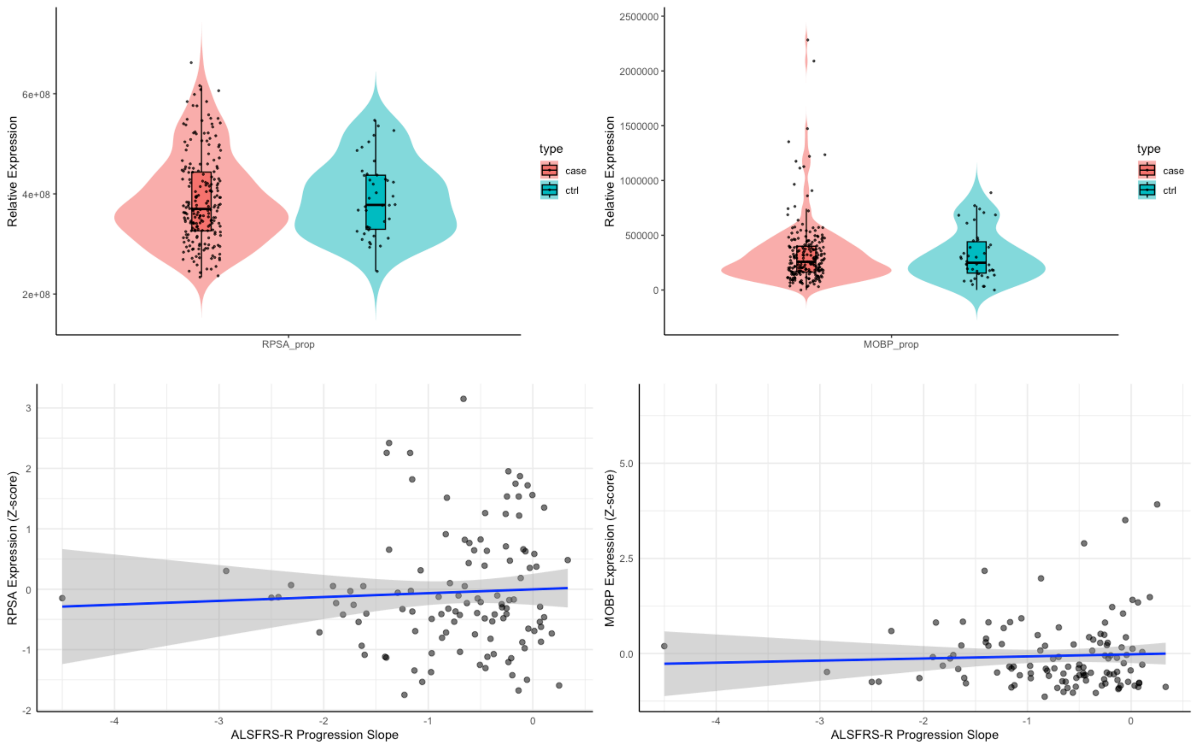


**Supplementary Figure 28**. Induced pluripotent derived motor neuron (iPSC-MN) expression of *RPSA* and *MOBP* expression **A.** No difference in ALS case vs. control expression of *RPSA* and *MOBP* (reads per million transcripts/TPM) (n=213 cases, n=42 controls) (age and sex as covariates, P-values 0.98 and 0.84 respectively). **B.** No association with level of expression and ALS progression rate (n=139) (age, sex, onset location as covariates, P-values were 0.89 and 0.84 respectively)

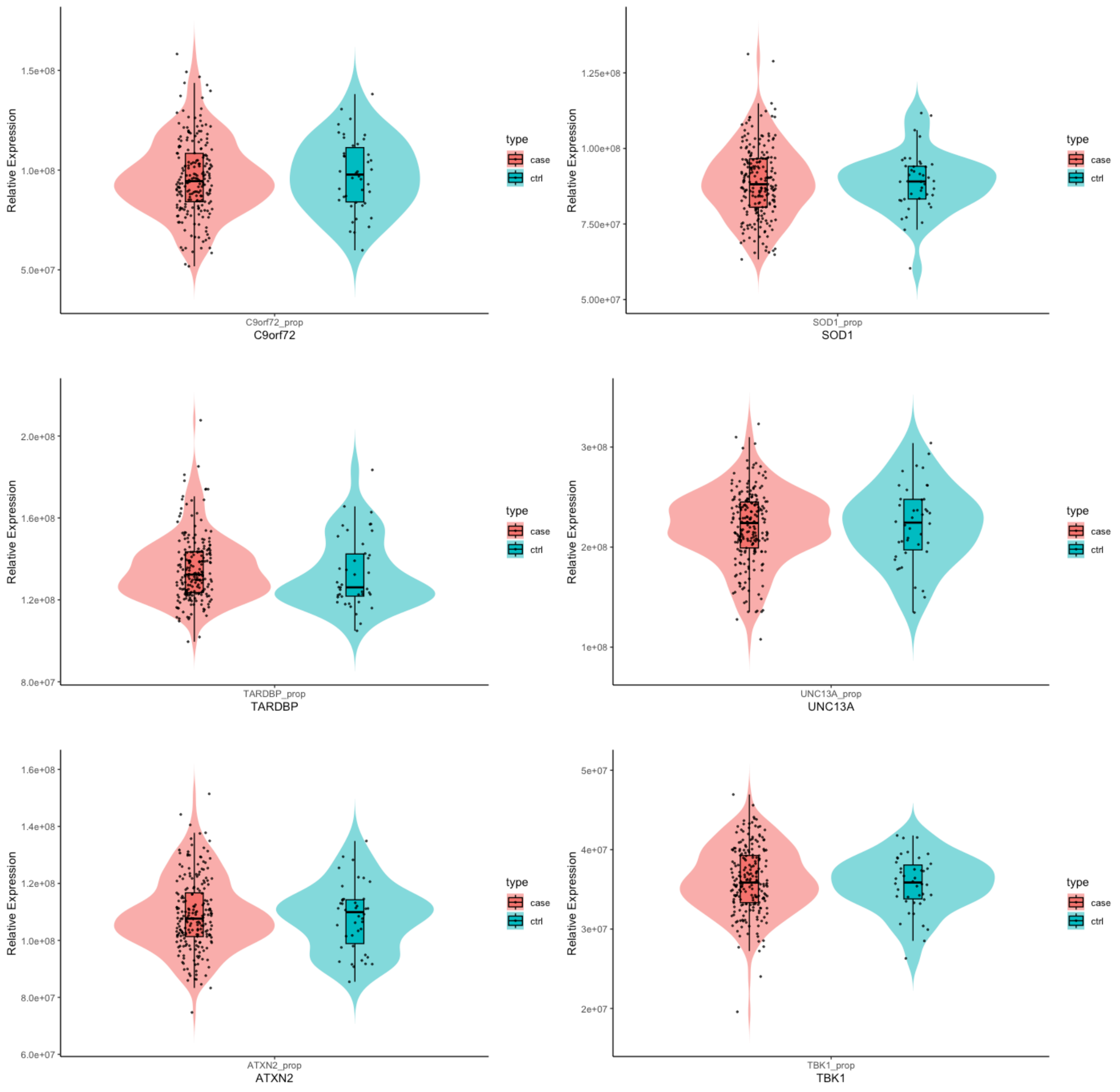


**Supplementary Figure 29**. Mendelian and risk gene expression in induced pluripotent derived motor neurons **A.** No difference in ALS case vs. control expression of *C9orf72 (P=0.62), SOD1 (P=0.34), TARDBP (P=0.80), UNC13A (P=0.84), ATXN2 (P=0.47)* and *TBK1 (P=0.70)* (reads per million transcripts/TPM) (n=213 cases, n=42 controls). Age and sex were included in the model as covariates.

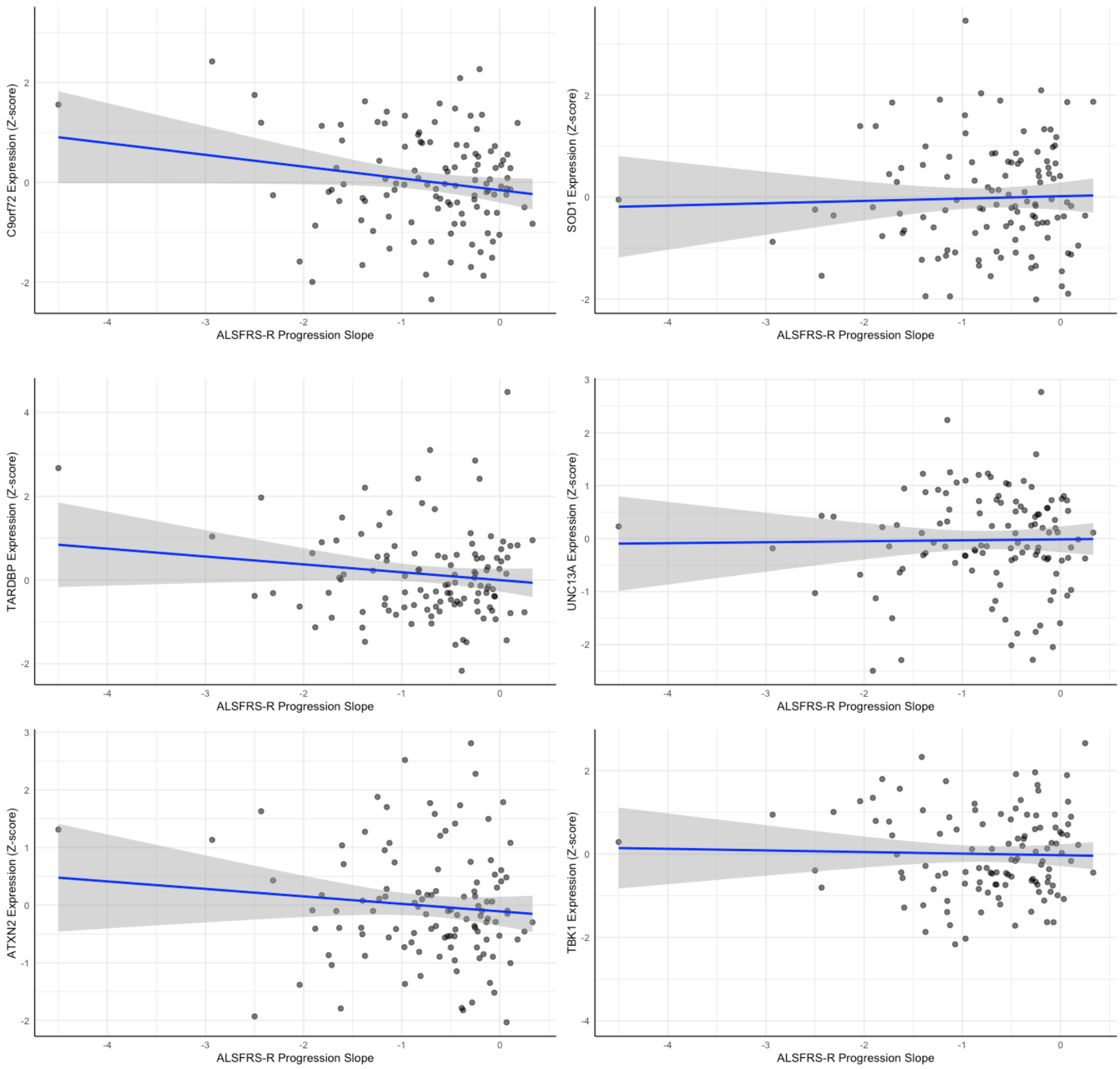


**Supplementary Figure 30**. Mendelian and risk gene expression in induced pluripotent derived motor neurons in ALS cases **A.** No association with level of expression of *C9orf72 (P=0.14), SOD1 (P=0.58), TARDBP (P=0.051, estimate=-0.26+- 0.13 SE), UNC13A (P=0.14), ATXN2 (P=0.41)* and *TBK1 (P=0.53)* and ALS progression rate (n=139). Age, sex, onset location were covariates.

```
CLUSTAL Ω (1.2.4) multiple sequence alignment

Hsa      MSGALDVLQMKEEDVLKFLAAGTHLGGTNLDFQMEHYIYKRKSDGIYIINLKRTWEKLLL 60
Dre      MSGGLDVLQMKEEDVLKFLAAGTHLGGTNLDFQMEQYVYKRKSDGVYIINLKKTWEKLLL 60
         ***.*******************************:*:*******:******:*******
Hsa      AARAIVAIENPADVSVISSRNTGQRAVLKFAAATGATPIAGRFTPGTFTNQIQAAFWEPR 120
Dre      AARAIVAIENPADVCVISSRNTGQRAVLKFASATGSTTFAGRFTPGTFTNQIQAAFREPR 120
         **************.****************:***:* :***************** ***
Hsa      LLVVTDPRADHQPLTEASYVNLPTIALCNTDSPLRYVDIAIPCNNKGAHSVGLMWWMLAR 180
Dre      LLIVTDPRADHQPLTEASYVNIPTIALCNTDSPLRYVDIAIPCNNKGPHSVGLMWWMLAR 180
         **:******************:************************* ************
Hsa      EVLRMRGTISREHPWEVMPDLYFYRDPEEIEKEEQAAAEKAVTKEEFQGEWTAPSPEFTA 240
Dre      EVLRMRGTISREHPWEVMPDLYFYRDPEEIEKEEQAAAEKAVGKEEFQGEWTAPVPDFN- 239
         ****************************************** *********** *:*.
Hsa      TQPEVADWSEGVQVPSVPIQQFPT---------------EDWSAQPATEDWSAAPTAQAT 285
Dre      -QPEVADWSEGVQVPSVPIQQFPAGIEAPGKPAPAEVYAEDWSAQPATEDWSAAPTAQAG 298
          **********************:               ********************
Hsa      EWVGATTDWS 295
Dre      DWGGATADWS 308
         :* ***:***
```

270/295 amino acid identity (92%).

**Supplementary Figure 31**. Alignment of human versus zebrafish *RPSA* protein sequence, highlighting exceptional conservation between the two organisms.
